# Quality control and removal of technical variation of NMR metabolic biomarker data in ∼120,000 UK Biobank participants

**DOI:** 10.1101/2021.09.24.21264079

**Authors:** Scott C. Ritchie, Praveen Surendran, Savita Karthikeyan, Samuel A. Lambert, Thomas Bolton, Lisa Pennells, John Danesh, Emanuele Di Angelantonio, Adam S. Butterworth, Michael Inouye

## Abstract

Metabolic biomarker data quantified by nuclear magnetic resonance (NMR) spectroscopy has recently become available in UK Biobank. Here, we describe procedures for quality control and removal of technical variation for this biomarker data, comprising 249 circulating metabolites, lipids, and lipoprotein sub-fractions on approximately 121,000 participants. We identify and characterise technical and biological factors associated with individual biomarkers and find that linear effects on individual biomarkers can combine in a non-linear fashion for 61 composite biomarkers and 81 biomarker ratios. We create an R package, ukbnmr, for extracting and normalising the metabolic biomarker data, then use ukbnmr to remove unwanted variation from the UK Biobank data. We make available code for re-deriving the 61 composite biomarkers and 81 ratios, and for further derivation of 76 additional biomarker ratios of potential biological significance. Finally, we demonstrate that removal of technical variation leads to increased signal for genetic and epidemiological studies of the NMR metabolic biomarkers in UK Biobank.

## Introduction

High-throughput NMR spectroscopy has enabled rapid simultaneous quantification of lipids, lipoproteins, fatty acids, and low-molecular weight metabolites including amino acids, ketone bodies, and glycolysis metabolites from a single human blood plasma sample (Ala-Korpela et al., 2021; Würtz et al., 2017). Over the last decade, NMR metabolic biomarker data has been quantified in numerous cohorts each with thousands participants, helping derive new insights into the epidemiology and molecular pathogenesis of cardiovascular and metabolic diseases and examine the genetic determinants of these molecular risk factors and disease biomarkers (Soininen et al., 2015).

The emergence and increasing scale of biobanks containing hundreds of thousands to millions of samples promises to enable discovery of new insights into human health and disease. To date, biomarker quantification has been completed in 121,695 participants (Julkunen et al., 2021), which have recently been made available for download by UK Biobank. Molecular phenotype data in large sample sizes are typically subject to the effects of unwanted technical variation (e.g. batch effects) which can obscure true biological effects and/or introduce false positive associations. Thus, a typical first step in any analysis is to identify and remove sources of unwanted variation from raw data. Here, we report additional quality control (QC) procedures to remove effects of technical variation on biomarker concentrations, which we make available as a resource to the community via an R package, ukbnmr.

## Results

### Summary of NMR metabolic biomarker data available in UK Biobank

UK Biobank is a deeply phenotyped cohort of approximately 500,000 thousand adults (Bycroft et al., 2018; Sudlow et al., 2015). Presently, NMR metabolic biomarkers have been quantified in approximately one third of randomly selected participants (**Methods**) (Julkunen et al., 2021). After sample QC (**Methods**), NMR metabolic biomarker data were available for 121,657 UK Biobank participants: 117,994 at baseline assessment (2006–2010) and 5,139 at first repeat assessment (2012–2013). The NMR metabolic biomarker data comprises absolute concentrations of 168 biomarkers along with 81 biomarker ratios (Würtz et al., 2017). These primarily consist of lipids and lipoprotein sub-fractions (81%) but also include fatty acids, amino acids, ketone bodies, glycolysis related metabolites, and others. An overview of available biomarkers and ratios are provided in **Figure 1**, and details are provided in **Table S1**.

**Figure 1:**
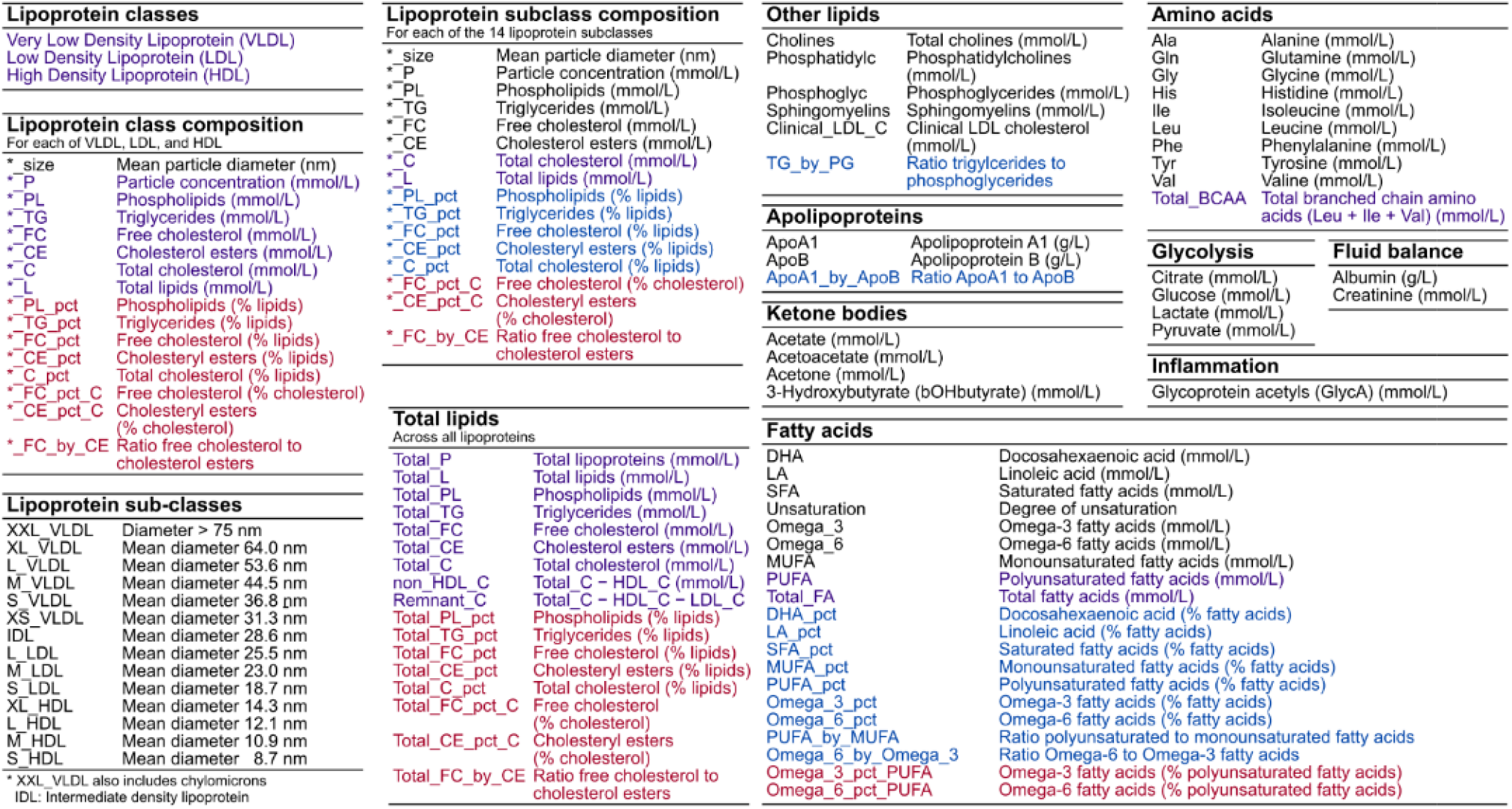
Summary of NMR metabolic biomarkers in UK Biobank. Short variable names, descriptions, and units for the 249 biomarkers and ratios available in the raw data as well as the 76 additional biomarker ratios derived in this study (shown in red). Biomarkers are grouped by type, given in each table heading. The 107 biomarkers in black are those which cannot be derived from any combination of other biomarkers. The 61 biomarkers in purple are the composite biomarkers that can be derived by summing two or more of the 107 non-derived biomarkers. Shown in blue are the 81 biomarker ratios available in the raw data, which can be derived from the 168 non-derived or composite biomarkers. Further details for each biomarker and ratio are provided in **Table S1** and formulae for deriving the composite biomarkers and ratios are provided in **Table S2**.

### Impact of technical variation

Data were recorded on a wide range of technical factors with potential to introduce unwanted variance in NMR metabolic biomarker measurements. These included shipping batch, 96-well plate, well position, aliquoting robot, and aliquot tip (UK Biobank), along with spectrometer and date and time stamps for each step in the biomarker quantification pipeline (Nightingale Health Plc.) (**Figure 2A, Methods**). **Figure 2B** shows the variance in raw biomarker concentrations explained by each technical covariate, and that this technical variance is removed by the QC procedures described below. Overall, most biomarkers were relatively robust to technical variation (**Figure 2B-C**), with median of 1.5% variance explained by the most correlated technical factor with each biomarker (**Figure 2C**).

**Figure 2:**
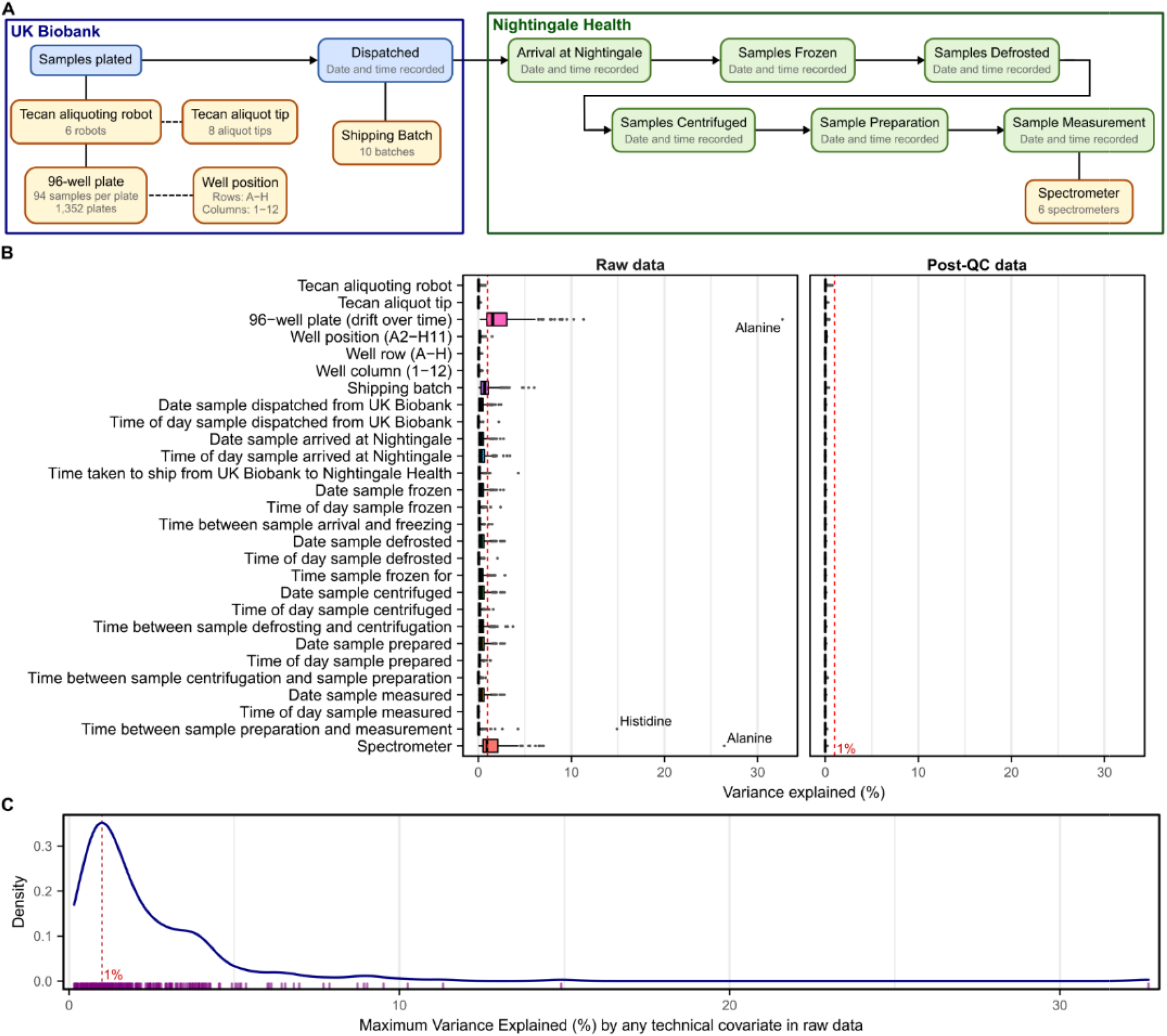
Technical covariates and their impact on biomarker concentrations. **A)** Schematic showing the technical covariates from the UK Biobank and Nightingale Health data processing pipelines (**Methods**). UK Biobank aliquoting robot and aliquot tip were randomised with respect to each other, with respect to 96-well plate, and with respect to position on 96 well plate. Plates and shipping batch were randomised with respect to spectrometer. **B)** Boxplots showing variance explained (**Methods**) across the 249 NMR metabolic biomarkers by each possible technical covariate in both the raw dataset and post-qc dataset. **C)** Density plot showing the maximum variance explained by any technical factor for each biomarker in the raw data. The rug plot below the density plot shows each of the 249 biomarkers.

There were 22 (of 249) biomarkers with at least 5% of variance explained by one or more technical factors, with strongest impacts observed for inter-spectrometer differences in biomarker concentrations, along with drift over time across different plates measured within each spectrometer (**Figure 2B**). Notably, 33% of the variance in alanine concentrations could be explained by drift over time within spectrometer (**Figure 2B, Figure 3A**) followed by 15% of the variance in histidine concentrations which could be explained by time between sample preparation and sample measurement (**Figure 2B, Figure 3B**). Further, inter-spectrometer differences in biomarker concentrations and drift over time within spectrometer were visually apparent for nearly all biomarkers (see extended diagnostic plots; **Data Availability**). Intra-plate variation was also visually apparent for some biomarkers (extended diagnostic plots; **Data Availability**), for example, we observed a consistent decrease in glycine concentrations from left to right across each plate (**Figure 3C**).

**Figure 3:**
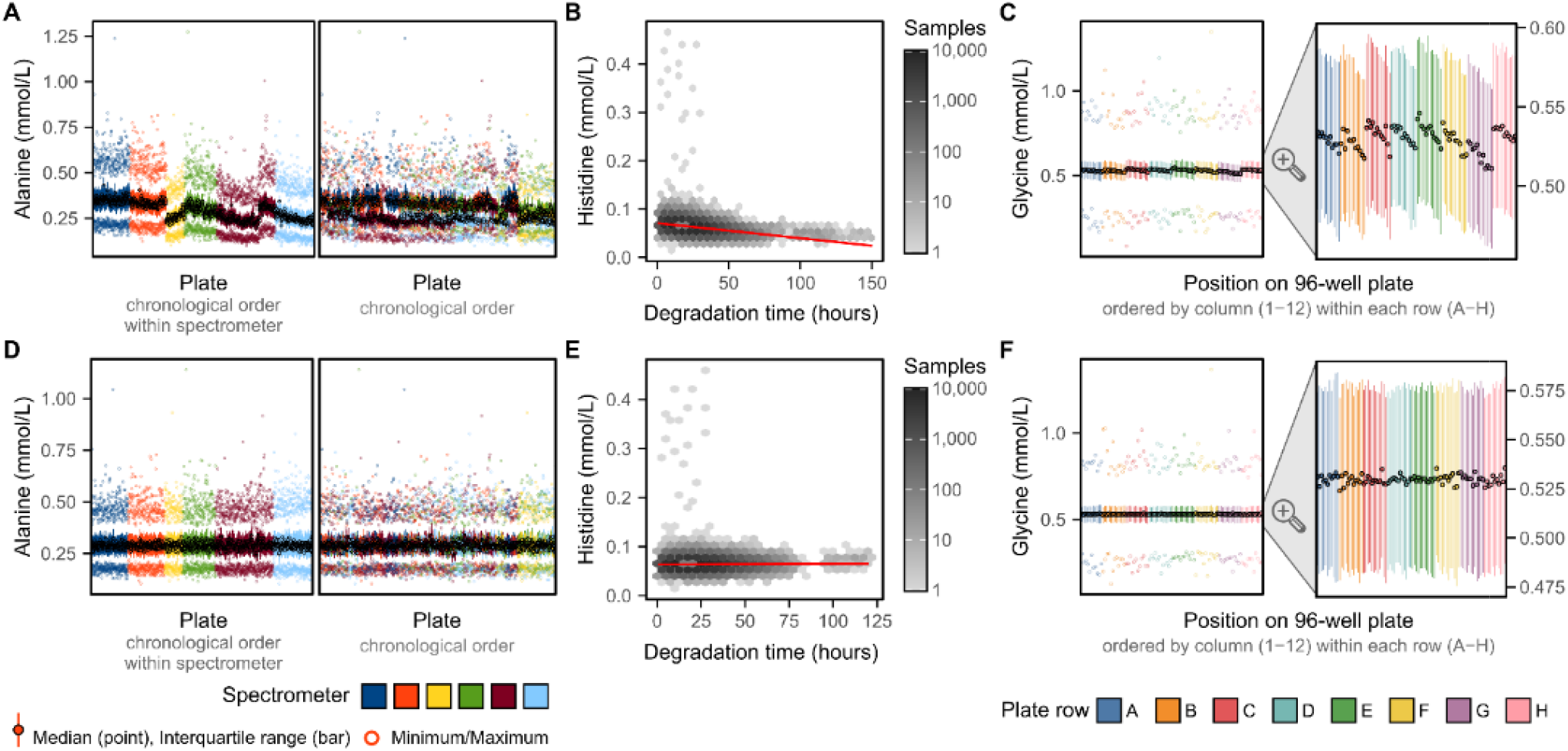
Strongest effects of technical covariates on biomarker concentrations. **A)** Inter-plate variation in alanine concentrations attributable to drift over time within spectrometer. Each plot shows a summary of alanine concentrations on each plate (minimum, maximum, median, and interquartile range). On the left plot, plates are ordered by date of plate measurement and grouped by spectrometer. On the right plot, plates are ordered by date of measurement without grouping by spectrometer. **B)** Histidine concentrations decrease as a function of sample degradation time (time between sample preparation and sample measurement). Hexagonal bins show sample counts on a log_10_ scale and red line shows the association (fit on all data points using robust linear regression). **C)** Glycine concentrations change as a function of plate position, with concentrations decreasing as plate column (1– 12) increase, and also systematically lower in plate row G. **D–F)** show the same as panels **A–C** after removal of technical variation.

### Removal of technical variation

Technical variation was subsequently removed using a multistep pipeline (**Methods**) detailed below. **Figure 4** gives a step-by-step example, for the NMR amino acid glycine, showing how concentrations and their relationship with technical covariates changes at each step.

**Figure 4:**
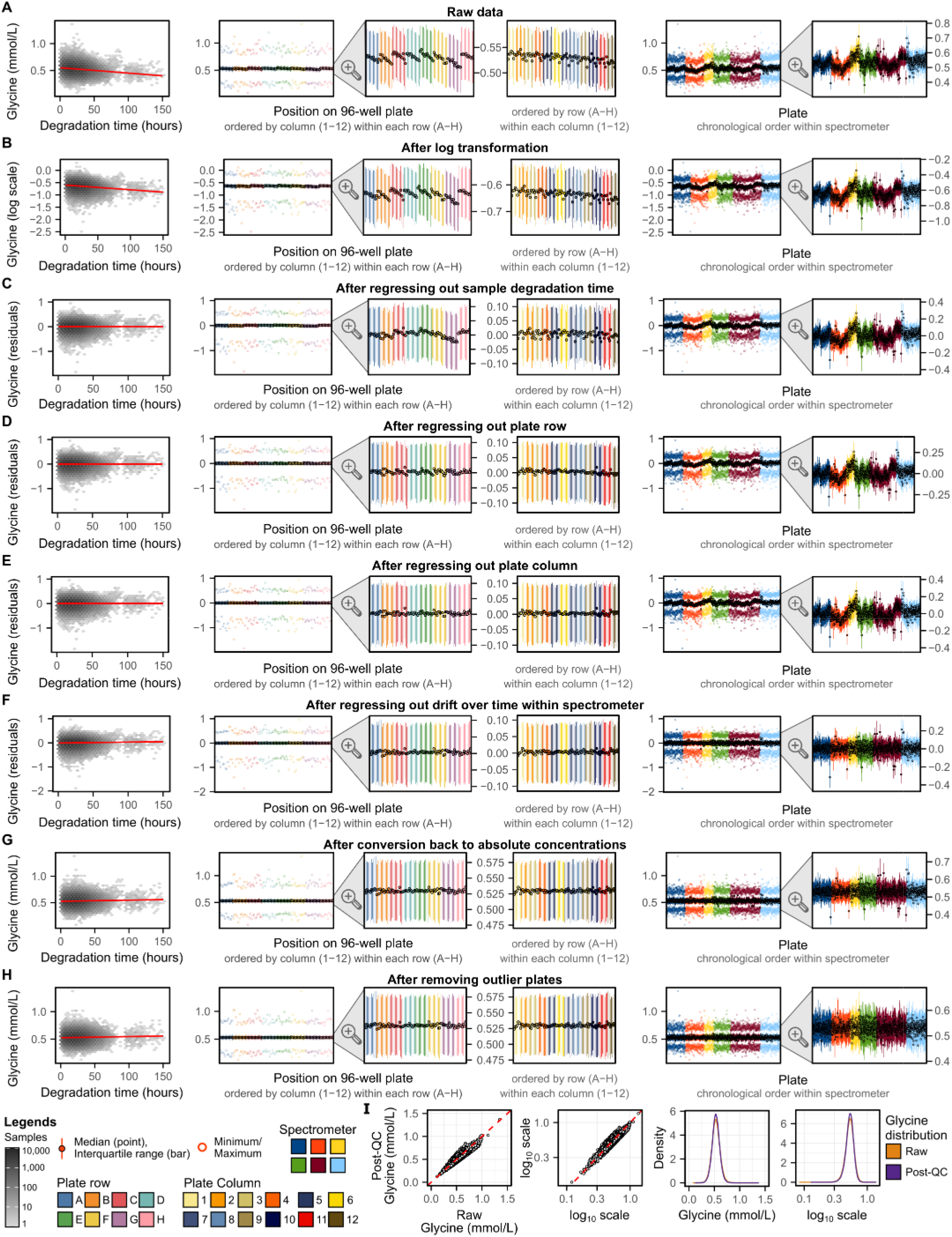
Step-by-step removal of technical variation for glycine concentrations. **A)** Glycine concentrations in the raw data and their relationship with technical covariates. **B-H)** Show how glycine levels and their relationship with technical covariates change after each step in the removal of technical variation pipeline (**Figure 1B, Methods**). In each of **A-H**, each plot shows from left to right: (1) relationship between glycine and sample degradation time (hours between sample preparation and sample measurement). Hexagonal bins show sample counts on a log_10_ scale and red line shows the association (fit on all data points using robust linear regression). (2) Summary of glycine levels on each well across all plates (minimum, maximum, median, and interquartile range). Wells are grouped and coloured by plate row (A–H) and within each row ordered by plate column (1–12). (3) A zoomed in view showing just the interquartile range and median for each well. (4) Alternative grouping of the zoomed in which wells are grouped and coloured by plate column (1–12) and within each column ordered by row (A–H). (5) Summary of glycine levels on each plate. Plates are grouped and coloured by spectrometer and within each spectrometer ordered by measurement date. (6) A zoomed in view showing just the interquartile range and median for each plate. **I)** Compares glycine concentrations before and after removal of technical variation. From left to right: (1) glycine concentrations in the raw data (x-axis) vs. glycine concentrations after removal of technical variation (y-axis) in mmol/L units, and (2) also shown on log_10_ scale (axis tick labels given in mmol/L units). (3) Distribution of Glycine concentrations in mmol/L units, and (4) also shown on log_10_ scale (axis tick labels given in mmol/L units). Plots showing how concentrations of all biomarkers change at each step and their relationship to technical covariates can be viewed on FigShare at doi: <u>10.6084/m9.figshare.16671208</u>.

A key motivating consideration at each step was to ensure adjustment for categorical technical factors did not break the samples into very small groups with potential to be non-random with respect to biological factors. For example, a simple approach to remove inter-plate variation would be to median normalise each plate. However, with only 94 samples measured per plate, this approach would likely remove not just technical variation but also biological variation of interest to downstream analysts. For example, allocation of participants to plates were not randomised with respect to participant sex (**Figure S1**), thus per-plate normalization would remove some sex-specific differences in metabolite concentrations, which may be of interest to downstream researchers.

With this consideration in mind, technical variation was removed through four sequential regressions (**Methods**). First, (1) biomarkers were regressed on time between sample preparation and sample measurement to remove any potential effects of sample degradation (**Figure 4C**). Next, (2) subsequent residuals were regressed on plate row (8 groups; rows A–H; **Figure 4D**) as visual differences between plate rows remained after removing effects of sample degradation time (**Figure 4C**). Third, (3) subsequent residuals were regressed on plate column (12 groups; columns 1–12; **Figure 4E**) as differences between plate columns were apparent for some biomarkers (**Figure S2**). Finally, (4) inter-plate variation due to drift over time within spectrometer was removed (**Figure 4F**) by binning plates into 10 groups by date within each of the 6 spectrometers (**Figure S3**) and regressing on bin as a categorical variable. At each of these steps robust linear regression (**Methods**) was used to fit robust linear regressions as we found linear regression susceptible to outliers and non-normality (**Figure S4**).

Prior to fitting these regressions, biomarker concentrations were log transformed (**Figure 4B**) so that a unit decrease in biomarker levels were equivalent to a unit increase (e.g. a halving and doubling both become a 0.69 unit change on the natural log scale). A small offset was applied for biomarkers with concentrations of 0 (**Table S3**). Subsequent to regressing out effects of technical variation, absolute biomarker concentrations were obtained (**Figure 4G**) from the regression residuals by rescaling their distribution to the raw data (**Methods, Figure S5**).

After removing most visible inter-plate variation (**Figure 4F-G**), we observed for many biomarkers several strong outlier plates (extended diagnostic plots; **Data Availability**). The strongest example, for albumin concentrations, is shown in **Figure 5A**. Stratification of albumin concentrations previously quantified by clinical biochemistry (Allen et al., 2020) according to the UK Biobank shipping plates used to send samples for NMR metabolite quantification supported a non-biological origin for the observed outlier plates (**Figure 5B**). Investigation of control samples placed on each plate by Nightingale Health Plc. to assess the NMR quantification process showed no deviation of control samples for outlier plates (**Figure 5C**), indicating the source of this technical variation was not due to the NMR quantification pipeline rather arose during the UK Biobank sample plating process. However, these outlier plates could not be adjusted for using any of the available technical covariates detailed in **Figure 2A**. We therefore systematically identified and removed these outlier plates (**Methods**) as the final step in the pipeline to remove unwanted technical variation (**Figure 4H**). Across biomarkers, these accounted for a median of 9 plates (0.66% of plates/samples), with maximum of 20 plates (1.5% plates/samples) for albumin and phosphoglycerides.

**Figure 5:**
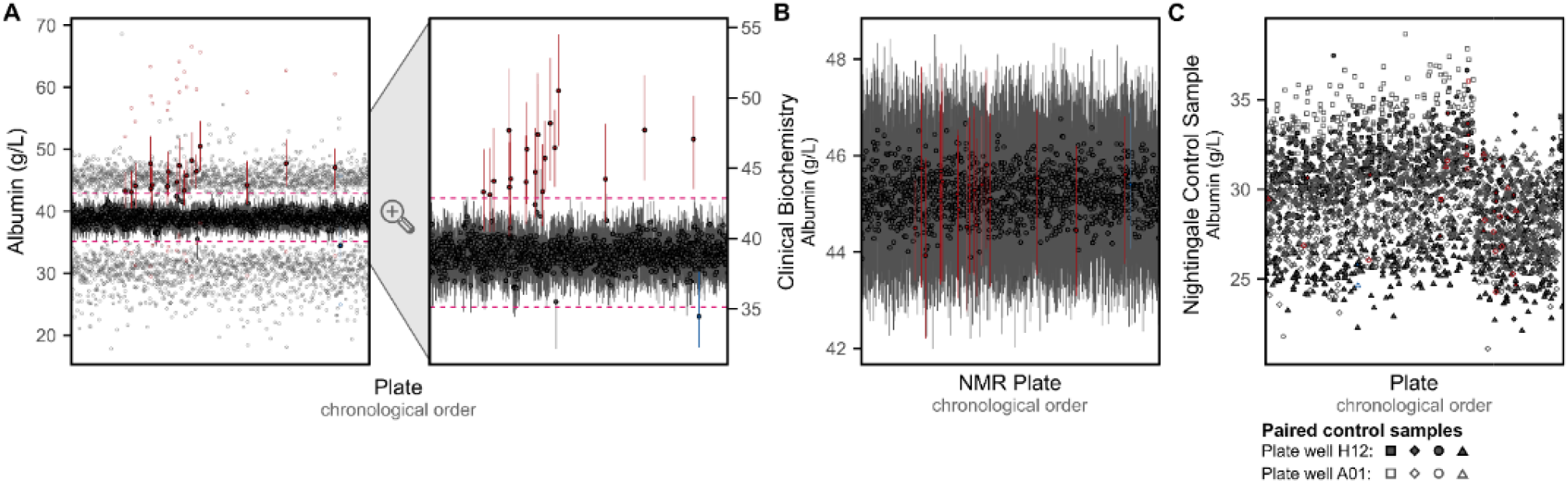
Outlier plates driven by unexplained technical variation. **A)** Summary of albumin concentrations on each UK Biobank shipping plate after removal of technical variation. For each plate, the median, interquartile range, minimum, and maximum values are shown on the left plot, with the second plot on the right showing just the median and interquartile range for each plate. Plates identified as outliers (**Methods**) are shown in red or blue. The horizontal pink dashed line show the limits above or below which plates were tagged as outliers based on their median values. **B)** Median and interquartile range of albumin concentrations previously measured by clinical biochemistry (Allen et al., 2020) when grouped by UK Biobank shipping plate used to send samples for NMR metabolite biomarker quantification. Plates that were identified as outliers in panel **A** are shown in red and blue. Of the 121,758 UK Biobank participants with NMR metabolite biomarkers, 107,283 also had albumin concentrations quantified by clinical biochemistry. **C)** Albumin concentrations quantified from control samples placed on plate wells A01 and H12 by Nightingale Health Plc. for internal quality checks in their NMR metabolite biomarker quantification pipeline. Four sets of paired control samples were used across all 1,352 plates. Note inter-plate and inter-spectrometer variation has not been removed from internal control samples here. Plates that were identified as outliers in panel **A** are shown in red and blue.

### Unwanted variation can combine in non-linear ways on composite biomarkers

Among the 249 NMR metabolite biomarkers available to download from UK Biobank, 81 were derived ratios, 61 were composite biomarkers, derivable as sums of two or more biomarkers (**Figure S6**), and 107 were non-derivable from other biomarkers (**Figure 1, Table S1, Table S2**).

Notably, we found that directly adjusting composite biomarkers or biomarker ratios for technical covariates sometimes led to different post-QC concentrations or ratios than obtained by computing the biomarker from its adjusted composite parts (**Figure 6A**). In particular, technical covariates could have different or even opposing effects on biomarkers contributing to a composite biomarker (**Figure 6B**), which could combine in a non-linear fashion on the composite biomarker (**Figure 6C**). These differences are proportional to the effects of the adjusted covariates on each biomarker: we highlight that these differences are much larger when adjusting for age, sex, and body mass index (BMI) (**Figure 6D**), which have much larger impacts on biomarker concentrations than technical covariates (**Figure 6E**).

**Figure 6:**
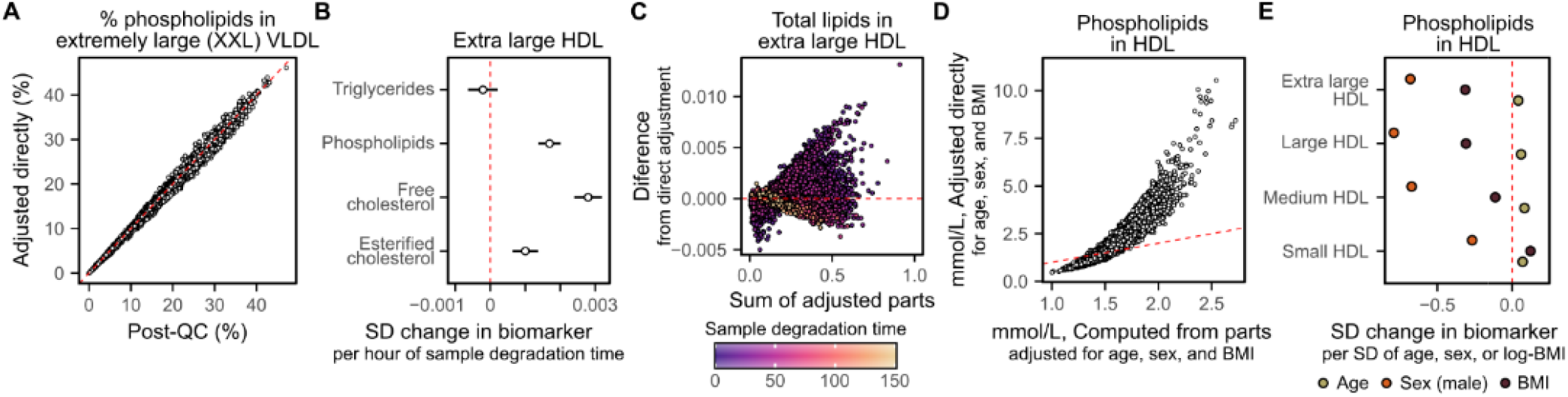
Composite biomarkers and ratios should be re-derived after removal of unwanted variation. **A)** Adjusting the % phospholipids in XXL VLDL for technical covariates (y-axes) leads to different values than computing % phospholipids in XXL VLDL from its component biomarkers (phospholipids in XXL VLDL over total lipids in XXL VLDL, where total lipids are the sum of free cholesterol, esterified cholesterol, phospholipids, and triglycerides) after adjustment for technical covariates. **B,C**) Technical covariates can have different effects on components of composite biomarkers that combine in non-linear ways. **B)** Shows for each of the components of total lipids in XL HDL the association from robust linear regression between sample degradation time and log biomarker concentrations in standard deviation units (horizontal bars show the 95% confidence intervals). **C)** Shows the difference between total lipids in XL HDL computed from its adjusted parts to total lipids in XL HDL directly adjusted for sample degradation time (y-axis) as a function of its concentration (x-axis) with points coloured by sample degradation time (hours). **D,E**) Shows how large differences can arise when adjusting for biological covariates with large effects on biomarker concentrations. **D)** Example showing non-linear differences between phospholipids in HDL computed from its adjusted parts (x-axis) to direct adjustment of phospholipids in HDL (y-axis) of the post-QC concentrations for age, sex, and BMI. **E)** Shows the association between age, sex, and BMI with concentrations of phospholipids in each HDL subclass from robust multivariable linear regression. Each point corresponds to the standard deviation change in the corresponding lipid per standard deviation increase in age, standard deviation increase in log transformed BMI, and when comparing male participants to female participants. 95% confidence intervals are smaller than the size of the shown points.

When removing the effects of technical variation above, we therefore recomputed the 61 composite biomarkers and 81 biomarker ratios after removing the effects of technical covariates from the 107 non-derivable biomarkers. We make available code for re-deriving these biomarkers and ratios in the ukbnmr R package (**Code Availability**).

### Derivation of additional biomarker ratios

We further derived 76 additional biomarker ratios not available in the original data (**Figure 1, Table S1, Table S2**), and make available code for deriving these additional biomarker ratios in the ukbnmr R package (**Code Availability**).

First, we derived 20 lipid fractions that are available for the 14 lipoprotein sub-classes but not for the lipoprotein classes and total serum. For each of the 14 lipoprotein sub-classes, the NMR metabolite biomarker data includes percentages of total lipids comprised of: (1) phospholipids, (2) triglycerides, (3) free cholesterol, (4) cholesteryl esters, and (5) total cholesterol (**Figure 1**). Here, we additionally derive these percentages for the three lipoprotein classes: very low density lipoprotein (VLDL), low density lipoprotein (LDL), and high density lipoprotein (HDL), as well as for total lipids in serum (**Figure 1**).

Next, we note that total cholesterol is composed of the sum of free cholesterol and esterified cholesterol (**Table S2**), thus derive cholesterol fractions (percentages of cholesterol made up of free cholesterol and esterified cholesterol) for total serum, the three lipoprotein classes, and 14 lipoprotein sub-classes (**Figure 1**). We also derived the ratio of free cholesterol to cholesteryl esters as there is some evidence that this ratio may be a determinant of lipoprotein atherogenicity (Bagheri et al., 2018).

Finally, we note that polyunsaturated fatty acids are composed of the sum of omega-3 and omega-6 fatty acids (**Table S2**), thus derive the percentage of polyunsaturated fatty acids comprised of omega-3 and omega-6 fatty acids (**Figure 1**).

### Quality control improves power for genetic and epidemiological studies

Next, we examined whether removal of technical variation impacted or improved power for genetic and epidemiological studies (**Methods**).

We performed genome-wide association analyses (GWAS) (**Methods**) for raw and post-QC concentrations of alanine, the biomarker most strongly affected by technical variation (**Figure 2B**), and albumin, the biomarker most strongly impacted by outlier plates (**Figure 5A**). We observed an increase in power for genetic associations for both biomarkers when using their post-QC concentrations with technical variation removed (**Figure 7A, Figure S7, Table S4**).

**Figure 7:**
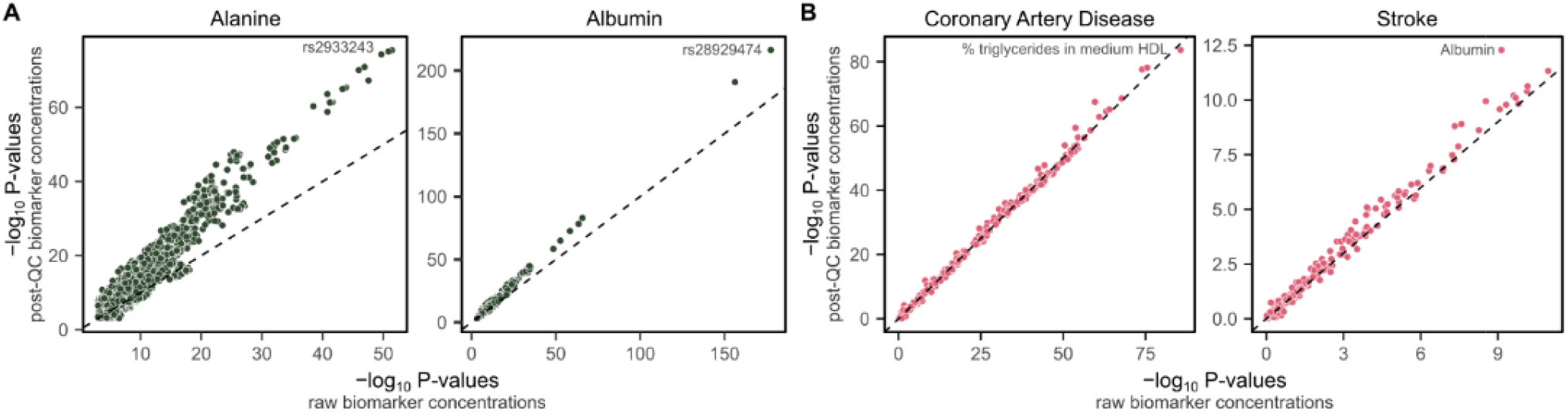
Adjustment for technical covariates improves power for genetic and epidemiological studies. **A)** Comparison of P-values from GWAS of 8.5 million common SNPs for alanine and albumin concentrations before and after removal of technical variation (**Methods**). The x-axes show –log_10_ P-values for associations between variants and raw concentrations, and y-axes show –log_10_ P-values for associations between variants and post-QC concentrations; after removal of technical variation. The dashed line on the diagonal shows y=x, where P-values are identical for both the raw and post-QC concentrations. The most strongly associated variant is annotated on each plot. Manhattan plots are shown in **Figure S7**. The number of genome-wide significant loci (P < 5×10^−8^) increased from 30 to 47 and 32 to 43 for alanine and albumin respectively when removing technical variation. **B)** Comparison of P-values from Cox proportional hazard models testing associations between incident coronary artery disease and incident stroke over 12.8 years of follow-up with raw and post-QC concentrations of the 249 NMR metabolite biomarkers (**Methods**). The x-axes show –log_10_ P-values for associations between raw biomarker concentrations and incident disease, and y-axes show –log_10_ P-values for associations between post-QC biomarker concentrations and incident disease. The dashed line on the diagonal shows y=x, where P-values are identical for both the raw and post-QC concentrations. The most strongly associated biomarker for each disease is annotated. Cox proportional hazard models were fit adjusting for age and sex. Participants with prevalent events or taking lipid lowering medication were excluded. Hazard ratios for all biomarkers are detailed in **Table S5**.

A comparatively modest difference in power was observed for biomarker association scans in Cox proportional hazards models for incident coronary artery disease and incident stroke (**Methods, Figure 7B, Table S5**). However, large differences in power are not expected in a biomarker association unless the biomarkers most strongly associated with the disease are also the ones most strongly impacted by technical variation. Notably, a large increase in power was observed for the association between albumin concentrations and incident stroke, which became the strongest association among the 249 biomarkers after removal of technical variation (**Figure 7B)**.

We also observed strong associations between the 76 additional biomarker ratios derived above with incident disease (**Table S5**), supporting their utility for future studies. For example, the percentage of LDL lipids composed of cholesterol was the biomarker with the strongest association with stroke (hazard ratio: 0.89, 95% confidence interval: 0.86–0.91, P-value: 2×10^−15^) above and beyond that of albumin (HR: 0.83, 95% CI: 0.79–0.87, P-value: 5×10^−13^).

### Characteristics of quality controlled NMR metabolite biomarker data

Finally, we explore and describe basic characteristics of the post-QC data of fundamental interest to downstream analysts in **Figure 8**. Specifically percentages of missing data across biomarkers (**Figure 8A**) and samples (**Figure 8B**), correlation between biomarkers (**Figure 8C**), and sources of inter-sample variation due to common physiological and environmental confounders (**Figure 8D-F**). In particular, we highlight that approximately 30% of the variation between biomarkers can be explained by a combination of sex, body mass index (BMI), and lipid lowering medication usage (**Figure 8E**), and that clustering of male and female participants is visible on PC1 and PC3 (**Figure 8F, Figure S8**). We also highlight that the structure of the data and relationships between biomarkers are largely unchanged from the raw data (**Figure S9**), with the exception of biomarker and sample missingness rates which are higher in the post-QC data (**Figure 8A-B**) due to the removal of outlier plates arising from unexplained technical variation (**Figure 5**).

**Figure 8:**
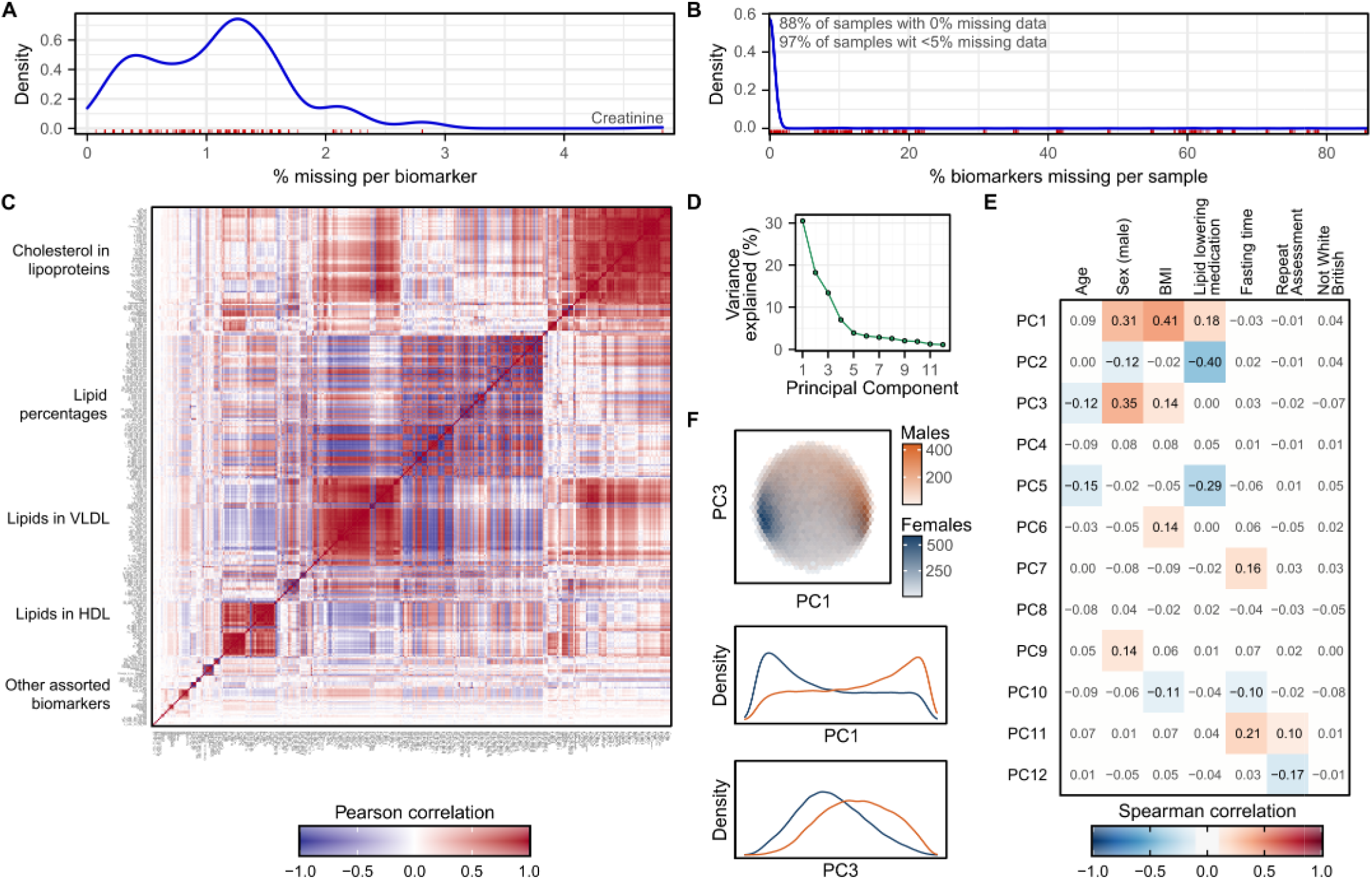
Characteristics of the NMR metabolite biomarker data after removal of technical variation. **A)** Density plot showing distribution of missingness across the 325 NMR metabolite biomarkers (including the 76 additionally derived biomarker ratios). The rug plot in red below the distribution shows the % of samples with missing data for each biomarker. **B)** Density plot showing distribution of missingness across the 123,023 samples. The rug plot in red below the distribution shows the % of missing biomarkers for each sample. **C)** Pairwise Pearson correlation coefficients between the 325 biomarkers. Biomarkers are ordered by hierarchical clustering based on their topological overlap dissimiliarity (Langfelder and Horvath, 2008) (**Methods**). Groups of correlated biomarkers are annotated. **D)** Principal components (PCs) explaining more than 1% of the variation in NMR metabolite levels between samples (**Methods**). **E)** Spearman correlation coefficients between the PCs and a selection of biological and environmental covariates. Heatmap cells are white and correlation coefficients are dulled where absolute value < 0.1. **F)** Separation of males and females by PC1 and PC3, the two PCs most strongly correlated with sex. The first plot shows hexagonal bins of samples, coloured by sex, with the two plots below showing density plots for PC1 and PC3 stratified by sex.

## Discussion

Technical variation is inherent to all laboratory measurements used for molecular phenotyping at biobank scale. Here, we comprehensively explored and document the sources and impacts of technical variation on the NMR metabolite biomarker data for ∼120,000 UK Biobank participants. We found that the vast majority of biomarkers are relatively robust to technical variation, but that a small subset are significantly impacted by inter-spectrometer variation, within-spectrometer drift over time, and intra-plate variation. We also found that a small number of plates are unexplained outliers of non-biological origin across a swathe of biomarkers. We subsequently developed a multi-step procedure for removing this unwanted variation, which we make available as part of the ukbnmr R package as a resource to the community.

We additionally highlight several important aspects of the NMR metabolite biomarker data analysts should be aware of.

First, there are major sources of biological variation that have systematic effects on biomarker concentrations between individuals. These include major differences in most biomarker concentrations depending on participant sex, body mass index, and use of lipid lowering medication, which are expected due to the predominantly lipid content of the biomarker data.

We also highlight the strong correlation structure between biomarkers, particularly among lipid concentrations, which are typical property for this type of data (Inouye et al., 2012; Würtz et al., 2015). This will directly impact choice of multiple testing strategies (Li and Ji, 2005), and can present challenges for multivariable modelling (Vatcheva et al., 2016), but can also be leveraged to increase power across multiple correlated traits (Nath et al., 2019). While analysts may reasonably be tempted to discard such highly correlated biomarkers, we highlight (Sliz et al., 2018) as a practical example of where highly correlated lipid fractions encoded different information. We also highlight (Ala-Korpela et al., 2021) as a recent dissection and discussion of the relative biological importance of various lipid ratios and percentages in lipoprotein sub-fractions, and (Soininen et al., 2015) as a review of findings from epidemiological and genetic studies for the NMR metabolic biomarkers more broadly. We would also highlight that filtering to non-derived biomarkers does not mitigate challenges introduced by collinearity, as many of the non-derived biomarkers are among those with extremely strong inter-correlations.

Analysts should also be aware that no filtering or treatment of outlier samples is performed by the ukbnmr R package when removing technical variation. Many biomarkers have extreme outliers whose treatment will need to be carefully considered for statistical modelling and many biomarkers have extremely non-normal distributions. One approach to handle these can be to winsorize biomarker distributions (either before or after log transformation) or to inverse rank normalize them.

Further, analysts should also be aware that Nightingale Health Plc has flagged some non-missing samples and biomarker measurements with QC flags (see UK Biobank fields #23651–23655 at https://biobank.ndph.ox.ac.uk/showcase/label.cgi?id=222 for sample QC flags and fields #23700–23948 at https://biobank.ndph.ox.ac.uk/showcase/label.cgi?id=221 for biomarker measurement QC flags). Sensitivity analysis to inclusion/exclusion of affected samples and measurements is recommended (see UK Biobank accompanying resources #3000 and #3004 at https://biobank.ndph.ox.ac.uk/showcase/label.cgi?id=220).

For some analysts, the post-QC dataset generated by our code in the ukbnmr R package may simply be a starting point for creating their own quality controlled datasets with further adjustment for unwanted biological variation, for example adjusting for age and sex for downstream GWAS. For these, we highlight that biomarker adjustment can introduce non-linear effects into biomarkers which are ratios, percentages, or otherwise composed of multiple other biomarkers. If creating new post-QC datasets, we recommend filtering to the 107 non-derived biomarkers and re-deriving the rest post-adjustment. We make publicly available methods for doing so in the R package ukbnmr. This caveat equally applies for anyone wishing to create new ratios or biomarker combinations. For example, here we additionally derive 76 lipid, cholesterol, and fatty acid percentages not present in the original Nightingale biomarker data.

In conclusion, we envision this manuscript and methods made available for removing technical variation in the ukbnmr R package will serve as a useful starting point for those who wish to use the NMR metabolite biomarker data available for ∼120,000 UK Biobank participants in their future studies.

## Methods

### NMR metabolite biomarker profiling of UK Biobank

UK Biobank is a cohort of approximately 500,000 individuals with deep phenotyping, imputed genotypes, and electronic health record linkage with written informed consent for health related research (Sudlow et al., 2015). Ethics approval was obtained from the North West Multi-Center Research Ethics Committee. The current analysis was approved under UK Biobank Projects 30418 and 7439. UK Biobank participants were members of the general UK population between 40 to 69 years of age identified and recruited through primary care lists and who accepted an invitation to attend one of 22 assessment centres across the UK between 2006 and 2010 (Sudlow et al., 2015). A subset of approximately 20,000 were selected for repeat assessment between 2012 and 2013 (Sudlow et al., 2015).

Absolute concentrations of 168 biomarkers and 81 biomarker ratios were quantified by NMR spectroscopy (Nightingale Health Plc.) from non-fasting plasma samples (UK Biobank aliquot 3) of 121,695 randomly selected UK Biobank participants as previously described (Julkunen et al., 2021). These included 117,981 participants at baseline assessment and 5,141 participants at repeat assessment, among which there were 1,427 participants with measurements at both time points.

Briefly, samples were randomly allocated to 96-well plates by UK Biobank and aliquoted in each well using one of six TECAN freedom EVO 150 robotic liquid handlers and one of eight 8 tips by UK Biobank. Aliquoting robot and aliquot tip were randomised with respect to each other, 96-well plate, and well position. Each plate contained a maximum of 94 samples from UK Biobank, with the remaining two wells (positions A01 and H12) reserved for internal control samples (Nightingale Health Plc.). Plates were randomly allocated to 10 batches and shipped to Nightingale Health (Helsinki, Finland). Plates were measured on one of six spectrometers (randomised with respect to UK Biobank shipping batch, aliquoting robot, and aliquot tip) by Nightingale Health. Further details on UK Biobank sample handling can be found in UK Biobank Resource 3000 (https://biobank.ndph.ox.ac.uk/showcase/showcase/docs/nmrm_companion_doc.pdf). Further details on the Nightingale Health NMR spectroscopy quantification pipeline are detailed in (Würtz et al., 2017) and details pertaining to UK Biobank detailed in (Julkunen et al., 2021).

### Sample quality control

Pre-release data made available to early access analysts, which was used to control for technical variation, differed in sample content from the raw data available to download from UK Biobank. In total 126,360 samples passed quality control in the pre-release data (**Supplementary Methods**). These included 6,359 blind duplicate samples: samples from participants sent by UK Biobank multiple times with differing sample identifiers for UK Biobank to assess the internal consistency of Nightingale Health’s NMR metabolite biomarker quantification pipeline. In total, 121,758 participants passed quality control: 118,047 with measurements at baseline assessment and 5,139 with measurements at first repeat assessment, including 1,428 participants with measurements at both timepoints.

For downstream analyses after removal of technical variation we filtered the pre-release data to the samples available for download from UK Biobank. Notably, the raw data available for download from UK Biobank included 37 samples that did not pass sample quality control in the pre-release data due to having insufficient sample material (**Supplementary Methods**). After removing these 37 samples, the raw and post-QC data used for downstream analyses contained 121,657 participants: 117,944 with measurements at baseline assessment and 5,139 with measurements at first repeat assessment, including 1,426 participants with measurements at both timepoints, with one sample per participant and timepoint.

### Variance explained by technical covariates

To estimate variance explained by each recorded technical covariate (**Figure 2B**), linear regression were fit for each biomarker and covariate separately and the model r^2^ obtained. Variance explained was estimated in both the raw and post QC biomarker data. For the post QC data, variance explained was estimated for both the 249 Nightingale biomarkers as well as the 76 additional biomarkers derived in the post-QC dataset.

For categorical variables (well position A02–H11, well row A–H, well column 1–12, spectrometer, shipping batch, aliquoting robot, and aliquot tip) the group with the largest sample size was used as the reference group. For categorical variables with missing data (aliquot tip, N=609 samples) the missing data were treated as a separate group, and not used as the reference group. For time of day events and date events (dispatch from UK Biobank, arrival at Nightingale Health, sample freezing, sample defrosting, sample centrifugation, sample preparation, and sample measurement) samples were split into 10 bins of equal duration and treated as a categorical variable (largest group as reference) to account for potentially non-linear effects over time. Durations between each pair of events were computed in hours (decimal) and treated as linear effects. To estimate variance explained by plate, plates were split into bins by spectrometer as described below in the removal of technical variation procedure, in this instance primarily due to the computational impracticality of fitting regression models on plate as a categorical variable (1,352 groups; regressions had not completed fitting for all biomarkers after 18 hours parallelized across 10 cores on high performance computing cluster).

### Removal of technical variation from known technical covariates

Raw biomarker data was filtered to 107 “non-derived” biomarkers: biomarkers that could not be expressed in terms of two or more other quantified biomarkers (**Table S1, Table S2**). Concentrations of these biomarkers were log transformed, with a small offset applied to biomarkers with 0 values (half the minimum non-zero value; **Table S3**).

Four sequential regressions were fit to remove the effects of known technical covariates (**Figure 4**). Log concentrations of each biomarker were (1) regressed on time between sample preparation and sample measurement using robust linear regression available through the MASS package in R (Huber, 2004; Venables and Ripley, 2002). Residuals were obtained, then (2) regressed on plate row as a categorical variable (8 groups; A–H) with robust linear regression using the row with the largest number of samples as the reference group (row D, N=16,155 samples). Residuals were obtained, then (3) regressed on plate column as a categorical variable (12 groups; 1–12) with robust linear regression using the column with the largest number of samples as the reference group (column 6, N=10,775 samples). Residuals we obtained, then (4) split into six groups by spectrometer. Plates within each spectrometer were ordered by measurement date, then split into 10 groups of approximately equal size (keeping plates measured on the same date in the same bin; **Figure S3**). Where measurement of samples on a plate occurred over multiple consecutive days, the plate measurement date was taken as the date on which the most samples were measured for that plate (N=487 plates with samples measured all on the same day, N=842 plates with samples measured over two consecutive days, N=22 plates with samples measured over three consecutive days, N=1 plate with samples measured over 4 consecutive days). Within each of the six spectrometer groups separately, robust linear regression was subsequently fit on plate measurement date bin as a categorical variable, using the bin with the largest number of samples as the reference group (N=1,306– 3,825 samples per reference group; **Figure S3**).

### Obtaining absolute concentrations for residuals

After removing the effects of known technical covariates from the 107 non-derived biomarkers, their residuals were converted back to absolute concentrations through the procedure described below (**Figure S5**).

The residuals from any regression are defined as the difference between the observed independent variable (e.g. biomarker concentrations) and the parameter estimated by the regression. A resulting key property is that the distribution of the residuals is centred on 0 for this estimated parameter. In the case of robust linear regression, this parameter is an estimate of the mean that is robust to outliers (Huber, 2004; Venables and Ripley, 2002). As a consequence of the way residuals are defined, their distribution can be scaled to match the distribution of the independent variable by giving it the same estimated mean. For robust linear regression, residuals can be returned to the same scale as the observed independent variable by estimating the mean of the observed independent variable using robust linear regression and adding it to the residuals.

When adjusting for technical covariates above, the independent variable was the log transformed raw concentrations of each biomarker. For each biomarker, we therefore fit a robust linear regression on the log transformed raw biomarker concentrations with just an intercept term to obtain an estimate of the mean robust to outliers. We then added this estimated mean to the residuals to return the residuals to the same scale as the log transformed raw concentrations. To return biomarkers to absolute concentrations, instead of log-concentrations, the exponent function was applied to inverse the log transformation. For biomarkers with concentrations of 0 in the raw data, the small offset applied prior to log transformation (**Table S3**) was also removed. Subsequently, some samples with concentrations of 0 in the raw data had negative concentrations very close to zero in the post-QC data; a small offset was applied to these biomarkers to ensure no negative concentrations (**Table S3**). In all cases this offset was at least one order of magnitude smaller than the smallest non-zero concentration, i.e. its impact on concentrations amounts to noise in numeric precision.

### Removal of outlier plates arising due to unexplained technical variation

Outlier plates were subsequently identified and removed for each of the 107 non-derived biomarkers (**Figure 4H, Figure 5**). To identify outlier plates, we modelled the distribution of plate medians for each biomarker as a normal distribution. For each biomarker, we computed the mean and standard deviation of plate medians across the 1,352 plates. Acceptable limits for plate medians (**Figure 5A**) were then set based on the limits of a theoretical normal distribution of 1,352 points. Plates were subsequently flagged and removed as outliers if their median concentration was greater than or less than 3.3744 standard deviations of mean of the 1,352 plate medians. A non-biological origin for outlier plates was confirmed by examining concentrations of biomarkers from previously quantified clinical biochemistry data (Allen et al., 2020) (**Figure 5B**).

### Derivation of composite biomarkers, ratios, and percentages

To create the final post-QC dataset, we subsequently recomputed the 61 composite biomarkers and 81 biomarker ratios available in the raw data from the 107 non-derived biomarkers using the ukbnmr R package (**Code Availability**, formulae in **Table S2**). We also computed 76 additional biomarker ratios not available in the original raw data (**Figure 1, Table S1, Table S2**).

### Comparison to direct adjustment

In **Figure 6A-C** we compared these computed post-QC biomarkers to those obtained by directly adjusting the biomarker for the technical covariates as described above. When doing so, percentage biomarkers (**Table S1**) were logit transformed rather than log transformed.

For **Figure 6D,E** we additionally created two datasets further adjusting for age, sex, and BMI: one in which the derived biomarkers were computed from the 107 non-derived biomarkers after adjustment, and one in which the derived biomarkers were directly adjusted. In both cases the starting point was the post-QC dataset described above, and robust linear regressions were fit for each biomarker on age, sex, and log transformed BMI. Post-QC biomarker concentrations were log or logit transformed as appropriate prior to fitting robust linear regressions, and residuals rescaled to absolute concentrations as described above.

### Genome-wide association study for alanine and albumin

Genome-wide association studies (GWASs) were performed for raw and post-QC alanine and albumin (**Figure 7A, Figure S7, Table S4**). GWAS were performed using the version 3 UK Biobank genotype data, which has been imputed to the 1000 genomes, UK10K, and Haplotype Reference Consortium panels (Bycroft et al., 2017; Loh et al., 2016) using human genome build GRCh37.

Participants were filtered to those of White British ancestry (from sample-QC file downloadable from UK Biobank in Resource 531) and filtered for relatedness (first- or second-degree relationships, kinship > 0.0884, from relatedness file downloadable from UK Biobank in Resource 531) (Manichaikul et al., 2010). For participants with measurements at both baseline and repeat assessment the measurement at baseline assessment was chosen. In total, 111,450 participants were included in the GWASs. Among these, 111,446 had non-missing data for raw alanine, 110,552 had non-missing data for post-QC alanine, 111,443 had non-missing data for raw albumin, and 109,803 had non-missing data for post-QC albumin.

GWAS were performed using generalized linear models using plink 2 (version 2.00a3LM AVX2 Intel; 2 Mar 2021) (Chang et al., 2015) on probabilistic dosage data extracted from UK Biobank’s Oxford BGEN format files (Band and Marchini, 2018). Associations were tested for 8,587,133 bi-allelic single nucleotide polymorphisms with minor allele frequency > 1% and imputation INFO score > 0.4. Associations were adjusted for age (UK Biobank field # 21003), sex (UK Biobank field # 22001), genotyping chip (UK Biobank in Resource 531), and 10 genotype PCs (UK Biobank in Resource 531) as covariates. Alanine and albumin concentrations were log-transformed prior to association testing and quantile normalized (along with covariates) by plink2.

GWAS peaks (**Figure S7, Table S4**) were identified by partitioning associations with P < 5×10^−8^ into 1,703 pre-defined linkage disequilibrium blocks that are approximately independent in populations of European ancestry (Berisa and Pickrell, 2016), and taking the SNP with the smallest P-value as the lead SNP for each peak. Lead SNPs were subsequently annotated using the Ensembl variant effect predictor (VEP) (McLaren et al., 2016), filtering to variants located within protein coding genes. Predictions of variant effects from PolyPhen-2 and SIFT were also obtained using the Ensembl VEP REST API (https://grch37.rest.ensembl.org/documentation/info/vep_id_post) (Adzhubei et al., 2010; Vaser et al., 2016). For lead SNPs not located within protein coding genes, the closest protein coding gene was identified using the annotables R package (Steinbaugh et al., 2017).

### Biomarker association scan for coronary artery disease and stroke

Cox proportional hazards models were fit using the R package survival (version 3.2-7) (Therneau and Grambsch, 2013) to test associations between raw and post-QC biomarker levels with incident coronary artery disease and incident stroke (**Figure 7B, Table S5**). Analysis was restricted to participants with NMR metabolite biomarker data at baseline assessment and who were not already taking lipid lowering medication (N=96,258 participants). Cox proportional hazard models were fit adjusting for age and sex (UK Biobank field #31), on standardised log transformed biomarker concentrations (logit transformation for percentages), and excluding prevalent case. Participant age, sex, and electronic health records were obtained under UK Biobank application number 7439. After excluding prevalent cases, for coronary artery disease analyses there were 95,790 samples including 2,717 incident cases, and for stroke analyses there were 95,623 samples including 1,366 cases. Follow-up in electronic health records was truncated at 1^st^ February 2020 to preclude any potential confounding from SARS-CoV2 exposure. The maximum and median follow-up time available in the electronic health records were 12.8 years and 10.9 years respectively.

Incident coronary artery disease was defined as the first occurring event of myocardial infarction (international classification of diseases (ICD) revision 10 codes I21–I24, and I25.2) or cardiovascular surgery (percutaneous transluminal coronary angioplasty: Office of Population Censuses and Surveys Classification of Surgical Operations and Procedures (OPSC) 4th revision codes K49, K50.1, and K75; or coronary artery bypass grafting OPSC-4 codes K40–K46). Prevalent CAD was similarly defined, with the addition of self-reported events (UKB field #6150 code “heart attack” and #20002 code 1075, UKB field #20004 code 1070, and UKB field #20004 codes 1095 and 1523). Retrospective hospital records included those using ICD revision 9 coding, for which ICD-9 codes 410–412 were used to identify previous hospitalisation with myocardial infarction.

Incident stroke was defined as the first occurring event of any stroke (ICD-10 codes I60–164) or fatal cerebrovascular events (ICD-10 codes I60–I69 and F10). Prevalent stroke was defined as any previous hospitalisation for stroke (ICD-10 codes I60–164 or ICD-9 codes 430, 431, 434, or 436) or self-reported stroke at UK Biobank assessment (UKB field #6150 code “Stroke” and #20002 codes 1081, 1086, 1491, or 1583).

### Hierarchical clustering of biomarker correlations

For **Figure 8C** pairwise Pearson correlations were computed between each pair of biomarkers in the post-QC dataset then clustered using the topological overlap dissimilarity distance metric available through the weighted gene coexpression network analysis (WGCNA) R package (version 1.69) (Langfelder and Horvath, 2008; Zhang and Horvath, 2005). Briefly, this distance metric computes the distance between biomarkers as a weighted sum of (1) the strength of correlation coefficient between a pair of biomarkers, and (2) the similarity of their correlation coefficients to all other biomarkers (Zhang and Horvath, 2005). To ensure strong negative correlations were clustered with strong positive correlations the absolute value of the pairwise Pearson correlation was taken prior to distance calculation, and a penalization exponent of 6 applied to shrink weak correlations towards zero as recommended in the package documentation (Langfelder and Horvath, 2008).

### Principal components analysis

For **Figure 8D**–**F, Figure S8**, and **Figure S9D**–**F**, principal components analysis (PCA) was performed across participants. For PCA, biomarker concentrations were first either logit transformed (percentages; **Table S1**) or log transformed (all other biomarkers) and standardised to have mean of 0 and unit variance. Small offsets were applied to biomarkers with 0 values to facilitate log transformation, and for percentages, small negative offsets were applied for those with values of 100%. Missing values were imputed as the biomarker mean (0) as PCA could not handle missing data. Ratios with infinite (x/0) or undefined (0/0) values were also set to missing prior to log transformation. The prcomp function in R was used for PCA.

To identify biological correlates of fitted principal components (PCs) (**Figure S8E, Figure S9E**), linked phenotype data was obtained for UK Biobank participants. Spearman correlation coefficients were fit between each PC and participant age, sex, BMI (UK Biobank field #21001), ethnicity (White British / other; UK Biobank field #21000), fasting time (UK Biobank field #74), and lipid lowering medication usage (UK Biobank fields #6153 and #6177). We also examined whether PCs correlated with UK Biobank assessment visit (repeat vs. baseline). Spearman correlation was used as several of biological and technical correlates were binary (yes/no) variables.

## Data Availability

All data described are available through UK Biobank subject to approval from the UK Biobank access committee. See https://www.ukbiobank.ac.uk/enable-your-research/apply-for-access for further details.
Extensive diagnostic plot showing the impact of technical variation and its removal on all biomarkers can be obtained from FigShare at doi: 10.6084/m9.figshare.16671208.

## Data Availability

All data described are available through UK Biobank subject to approval from the UK Biobank access committee. See https://www.ukbiobank.ac.uk/enable-your-research/apply-for-access for further details.

Extensive diagnostic plot showing the impact of technical variation and its removal on all biomarkers can be obtained from FigShare at doi: <u>10.6084/m9.figshare.16671208</u>.

## Code Availability

Code for removing technical variation from the NMR biomarker data and for re-computing composite biomarkers and ratios is available through the R package ukbnmr, which also provides tools for extracting and processing NMR metabolite biomarker data and associated quality control tags from UK Biobank. The ukbnmr R package and installation instructions can be found at https://github.com/sritchie73/ukbnmr/, and the package will be submitted to CRAN subsequent to peer review of this preprint.

Code underlying this paper are separately available at https://github.com/sritchie73/UKB_NMR_QC_paper/. This repository and specific release for this paper are permanently archived by Zenodo (European Organization for Nuclear Research and OpenAIRE, 2013) at doi: <u>10.5281/zenodo.5526029</u>.

## Acknowledgements

The authors are grateful to UK Biobank for access to data to undertake this study (Projects #30418 and #7439).

Nightingale Health Plc is acknowledged for early access to the UK Biobank NMR biomarker data and discussions regarding sources of experimental variation.

This work was performed using resources provided by the Cambridge Service for Data Driven Discovery (CSD3) operated by the University of Cambridge Research Computing Service (www.csd3.cam.ac.uk), provided by Dell EMC and Intel using Tier-2 funding from the Engineering and Physical Sciences Research Council (capital grant EP/P020259/1), and DiRAC funding from the Science and Technology Facilities Council (www.dirac.ac.uk).

S.C.R was funded by the National Institute for Health Research (NIHR) Cambridge BRC (BRC-1215-20014). P.S. was supported by a Rutherford Fund Fellowship from the Medical Research Council grant MR/S003746/1. S.A.L. is supported by a Canadian Institutes of Health Research postdoctoral fellowship (MFE-171279). T.B. was funded by the NIHR Blood and Transplant Research Unit in Donor Health and Genomics (NIHR BTRU-2014-10024). L.P. and S.K. were funded by a BHF Programme Grant (RG/18/13/33946). J.D. holds a British Heart Foundation Professorship and a NIHR Senior Investigator Award.

The funders had no role in study design, data collection and analysis, decision to publish, or preparation of the manuscript. The views expressed are those of the author(s) and not necessarily those of the NIHR or the Department of Health and Social Care.

