## Supplementary Information for "Quality control and removal of technical variation of NMR metabolic biomarker data in ∼120,000 UK Biobank participants"

#### Supplementary Figures

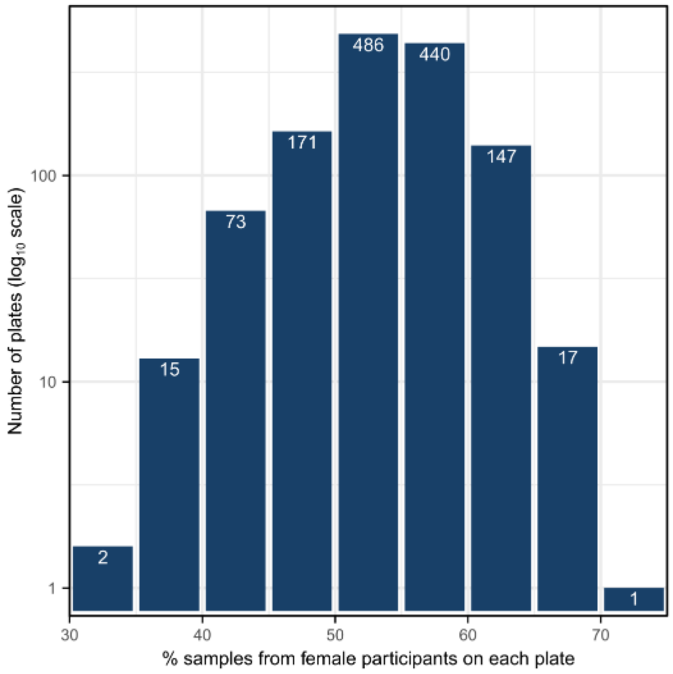

**Figure S1: Allocation of UKB participant to plate is not random with respect to participant sex.** Histogram shows the number of plates by percentage of samples from female participants in bins of 5%. The number of plates in each bin is annotated.

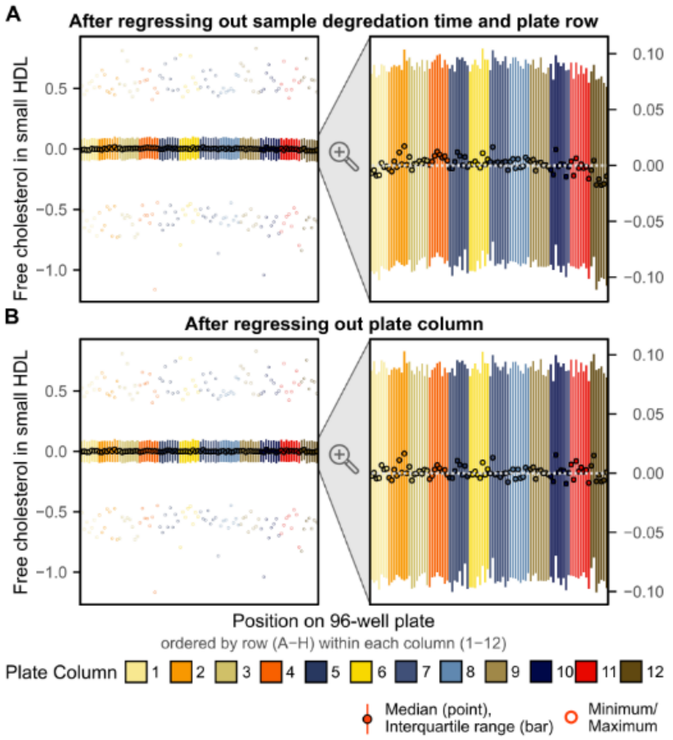

**Figure S2: Example of intra-plate variation across plate columns.**

**A)** Shows inter-column variation for free cholesterol in small HDL after regressing out sample degradation time and plate row. The left plot shows a summary of the regression residuals (minimum, maximum, interquartile range, and median) for each plate position grouped and coloured by plate column (1–12) and ordered within each column by plate row (A–H). A zoomed in view focusing on the interquartile range is shown on the right. A grey dotted line is overlaid at y=0, lower values can be seen for samples measured in columns on the outside of the plate (columns 1 and 12). **B)** Shows a summary of residuals for each plate position after regressing out plate column number as a categorical variable.

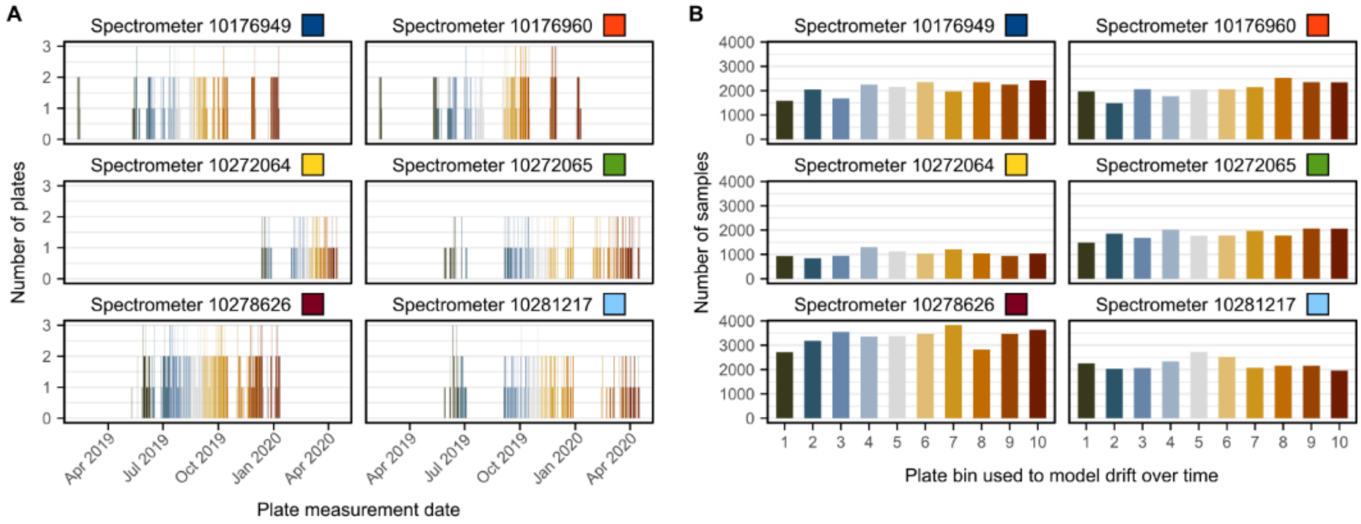

**Figure S3: Binning of plates within spectrometer to model drift over time.** For each of the six spectrometers, plates were ordered by measurement date, and split into 10 bins containing approximately equal number of plates (plates measured on the same date were allocated to the same bin). **A)** Shows the number of plates measured on each date, coloured by bin, with each plot corresponding to a single spectrometer. Colours next to each spectrometer indicate the colours used in **Figure 3D** and **Figure 4** when plotting summaries of biomarker concentrations on each plate. **B)** Shows the total number of samples measured on plates allocated to each bin. Within each spectrometer plate bin treated as a categorical variable and the bin with the largest number of samples was used as the reference group to regress out drift over time with spectrometer (**Methods**, **Figure S3F**).

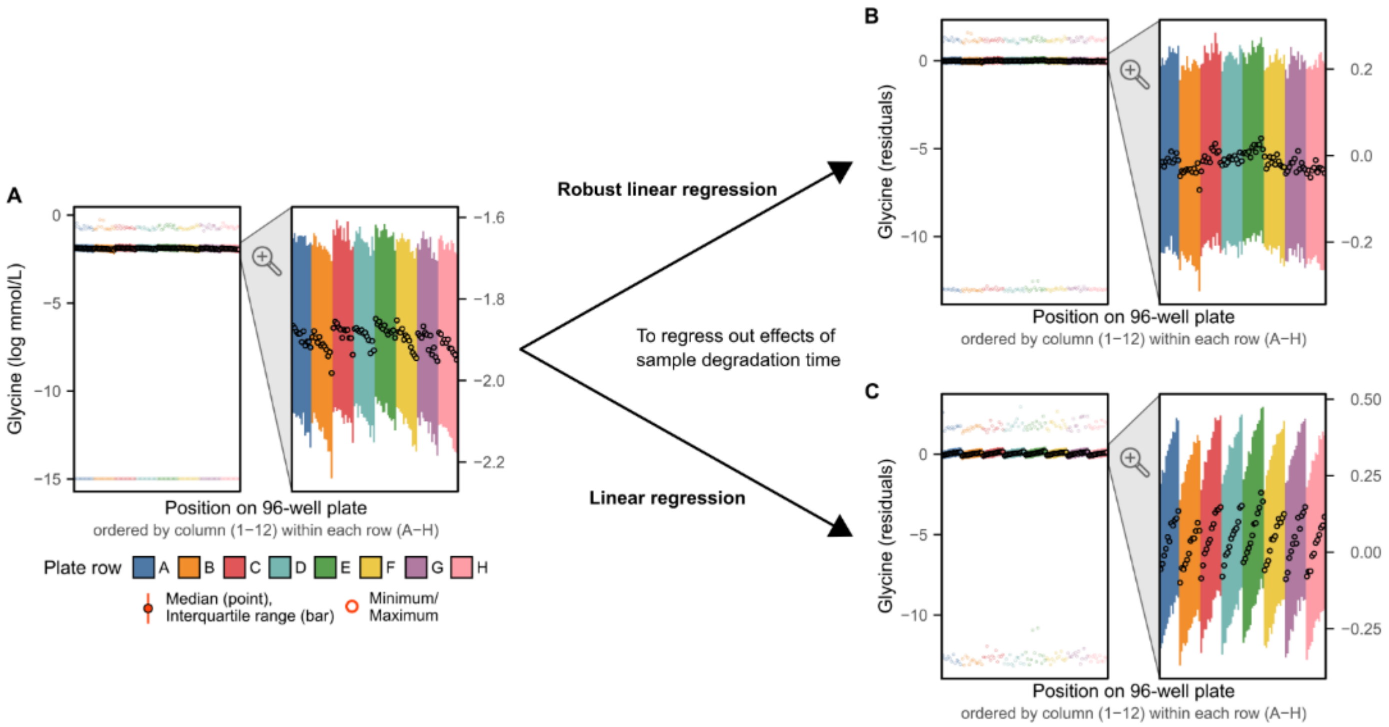

**Figure S4: Non-normality of biomarker concentrations requires robust linear regression.** Example comparing outcome of regressing glycine (raw values after log transformation; **A**) on time between sample preparation and sample measurement (sample degradation time) using (**B**) robust linear regression vs. (**C**) linear regression. Each plot shows a summary of glycine across all plates at each plate position (minimum, maximum, median, and interquartile range) with plate positions coloured by plate row (A–H), and within each row organised from left to right by plate column (1–12). Zoomed in plots focus on the interquartile range. In **A)** units are log mmol/L, while **B)** and **C)** show residuals from the respective regression.

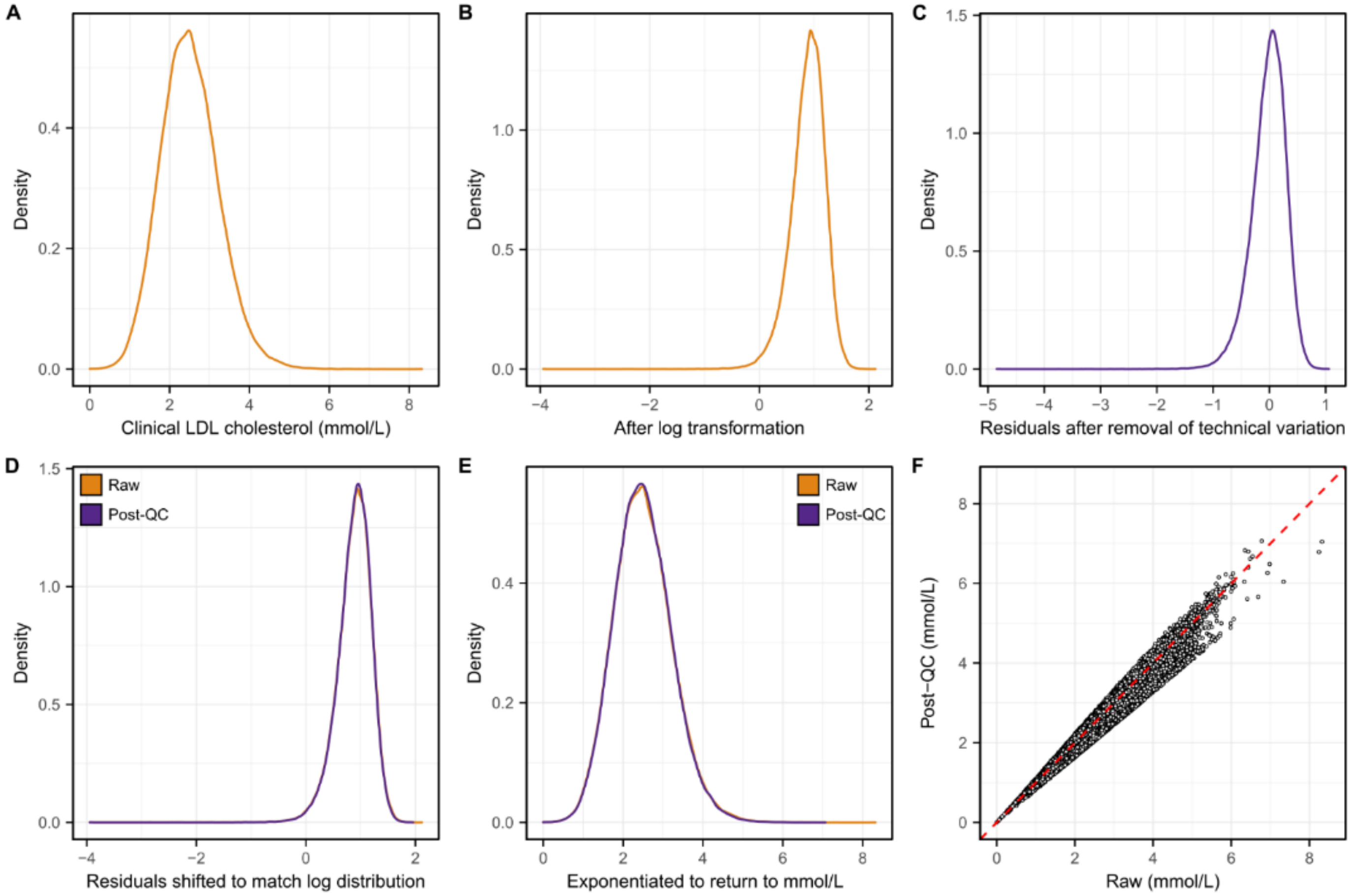

**Figure S5: Obtaining absolute concentrations after removal of technical variation.** Example given using concentrations of clinical low-density lipoprotein (LDL) cholesterol. **A)** Shows the distribution in the raw data. **B)** Shows the distribution after log transformation. As some samples had a concentration of 0 mmol/L, a small offset (half the minimum non-zero value: 0.0194; **Table S3**) was applied to all concentrations to enable these samples to be included after log transformation. **C)** Shows the distribution of the residuals after regressing out in four sequential robust linear regressions sample degradation time, then plate row, then plate column, the plate drift over time within spectrometer. **D)** Shows the distribution of residuals overlaid with the distribution of log raw concentrations from **Figure S8B** after shifting the residuals distribution to have the same median as the log raw distribution. **E)** Absolute concentrations are subsequently obtained by taking the exponent of the shifted residuals and removing the originally applied offset of 0.0194 (**Table S3**) from all samples. A final offset of $4.43\times10^{-5}$ (**Table S3**) is applied to ensure there are no negative concentrations. Here, the distribution of absolute concentrations after removal of technical variation is shown (purple) overlaid the distribution from the raw data shown in **Figure S8A**. **F)** Scatterplot comparing concentrations of clinical LDL cholesterol in the raw data (x-axis) to concentrations after removal of technical variation (y-axis). The red dashed line on the diagonal shows y=x, where concentrations fall if unchanged.

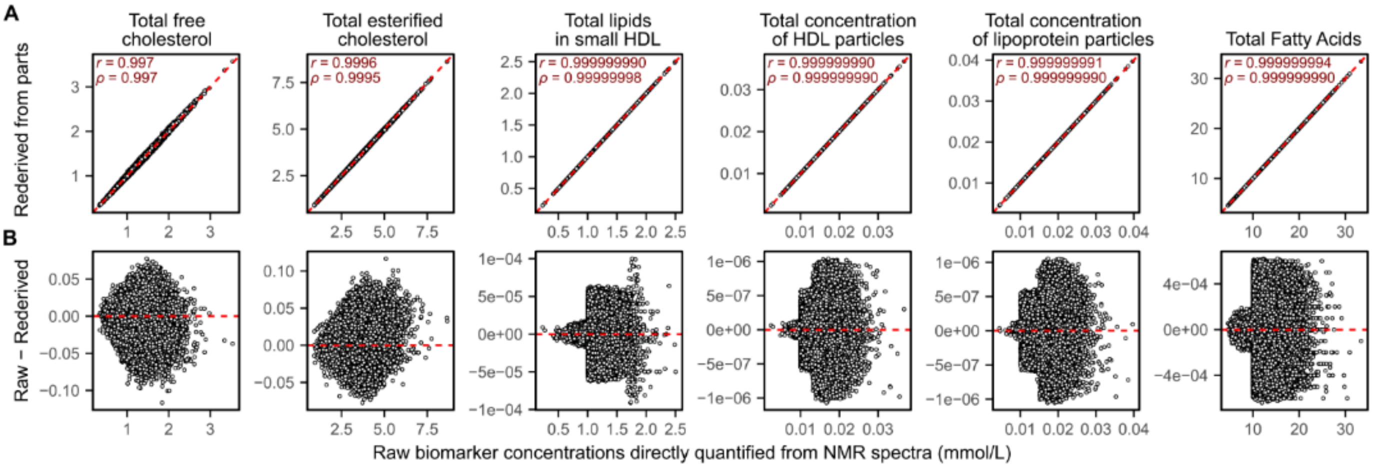

**Figure S6: Composite biomarkers can be re-derived without loss of information**

**A)** Compares raw biomarker concentrations (x-axes), quantified directly from the NMR spectra, to concentrations recomputed from the biomarker's parts (y-axes) for the six biomarkers most different (by Pearson and Spearman correlation) after re-derivation. The Pearson correlation coefficient (*r*) and Spearman rank correlation coefficient ($\rho$) between raw concentrations and recomputed concentrations are given in the top left of each plot. The dashed red line in each plot shows y=x. Formulae for each biomarker from parts are given in **Table S2**. For example, total free cholesterol can be computed be summing the concentrations of free cholesterol in the 14 lipoprotein subclasses. **B)** Shows the difference between raw and re-derived biomarker levels (y-axes) as a function of raw biomarker levels (x-axes), and that these differences are within numeric error for any given value of the biomarker.

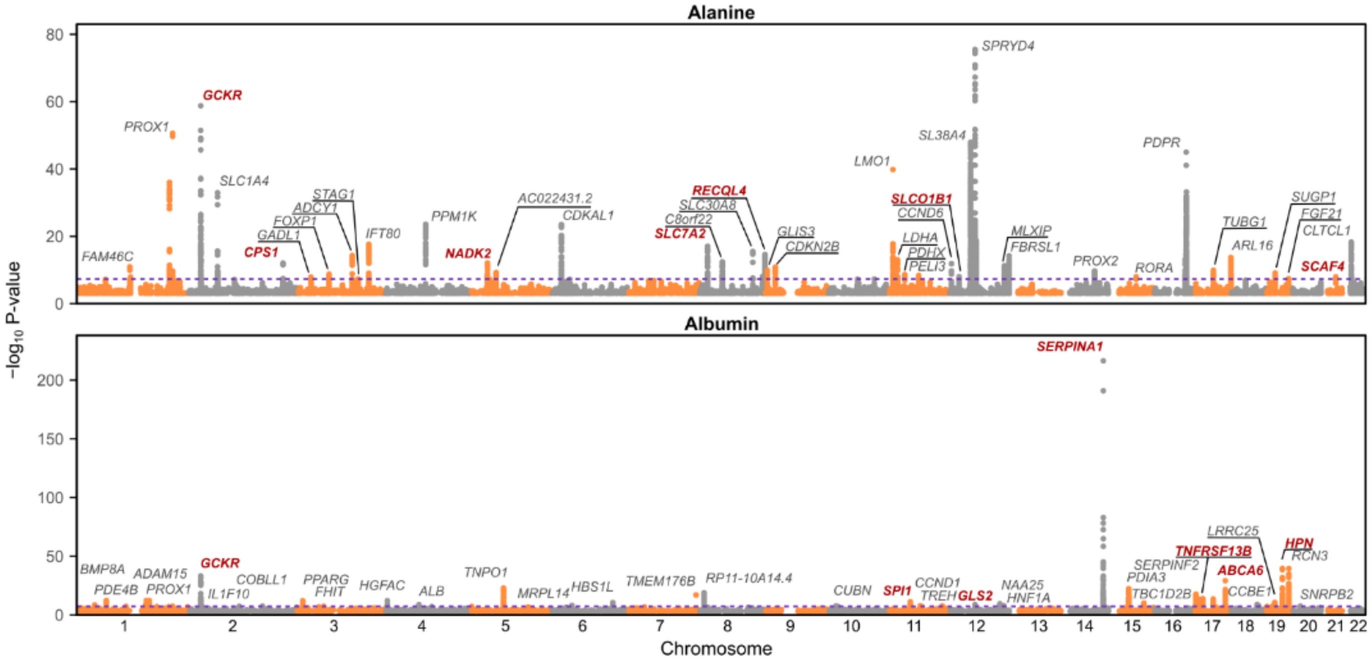

**Figure S7: Manhattan plots for GWAS of alanine and albumin after removing technical variation**
Manhattan plots show −log10 P-values from GWAS of 8.5 million common (frequency > 1%) SNPs (**Methods**). P-values greater than 0.001 are omitted. Peaks are annotated with their nearest protein coding gene. Genes in red indicate protein coding SNPs. Annotations for all peaks are detailed in **Table S4**.

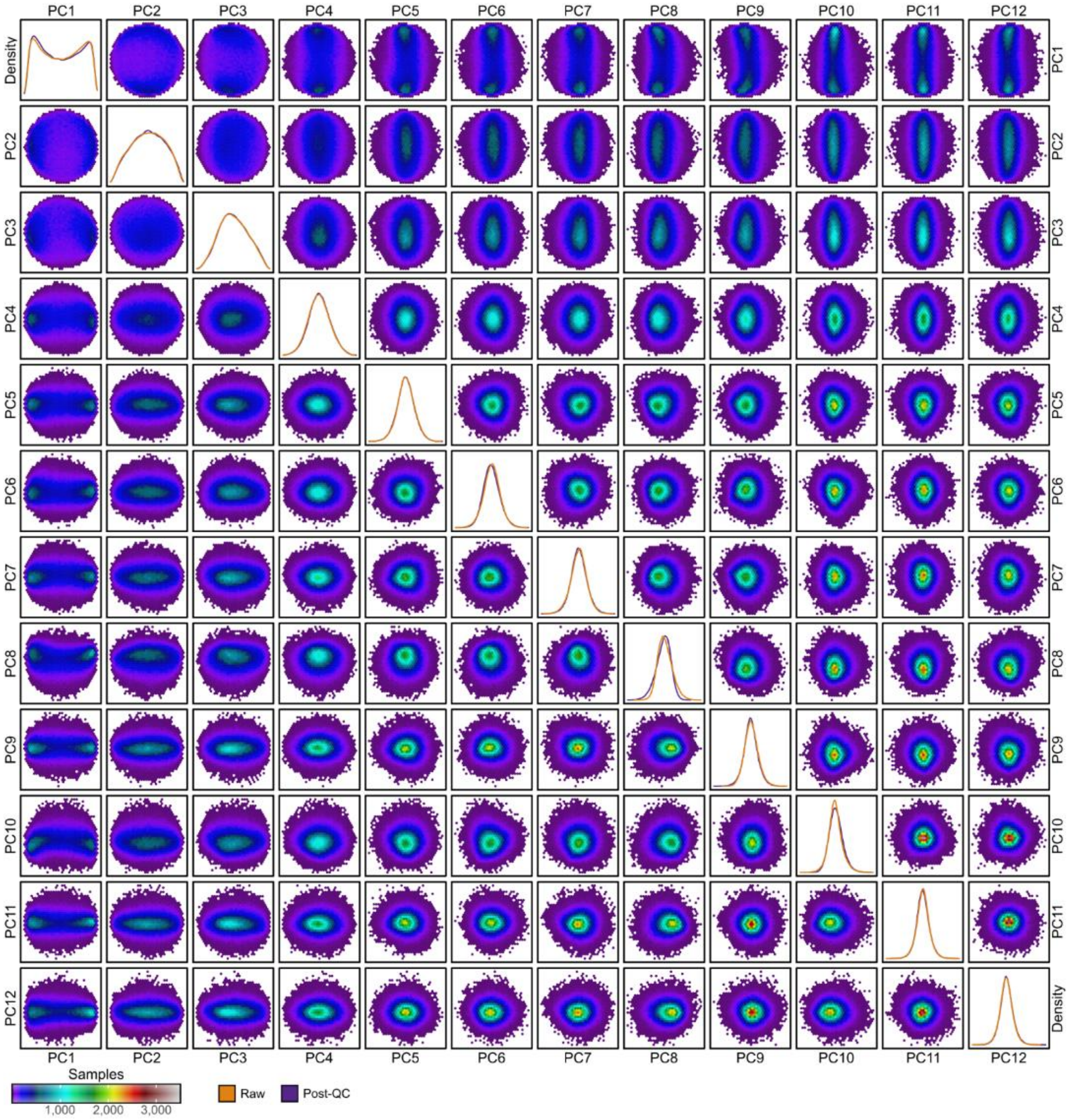

**Figure S8: Principal components explaining more than 1% of variation in samples.** Heatmaps show bivariate sample densities for each pair of the first 12 principal components (PCs) with each hexagonal cell coloured based on sample count. Heatmaps above the diagonal show the PCs computed from the raw data, and heatmaps below the diagonal show the PCs computed from the data after removal of technical variation (post-QC data). The diagonal shows density plots for the distributions of each PC in the raw and post-QC data.

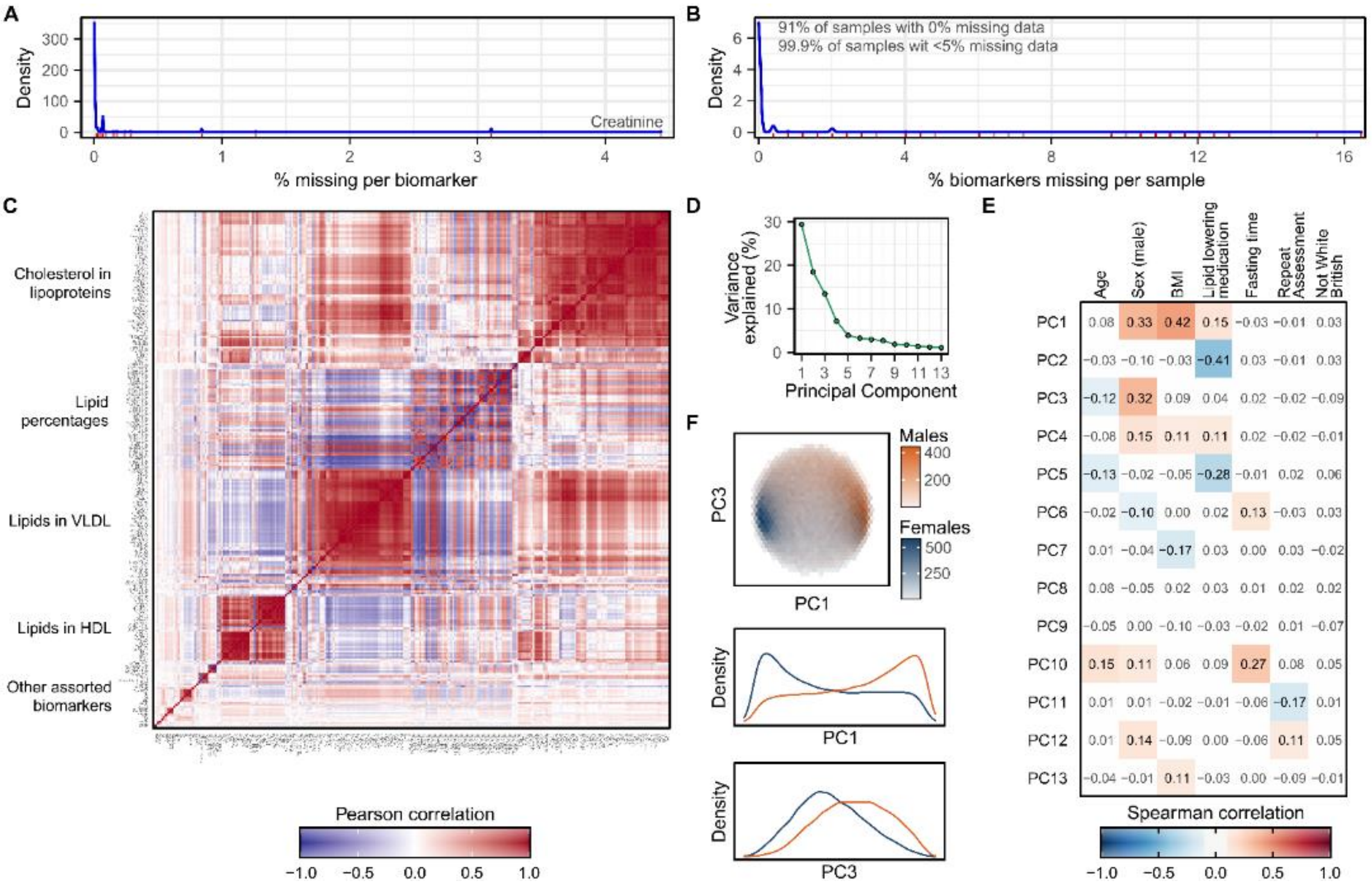

**Figure S9: Characteristics of the raw NMR metabolite biomarker data**

**A)** Density plot showing distribution of missingness across the 249 NMR metabolite biomarkers in the raw UK Biobank data. The rug plot in red below the distribution shows the % of samples with missing data for each biomarker. **B)** Density plot showing distribution of missingness across the 123,023 samples. The rug plot in red below the distribution shows the % of missing biomarkers for each sample. **C)** Pairwise Pearson correlation coefficients between the 249 biomarkers, shown in the same row and column order as in **Figure 8C**. Groups of correlated biomarkers are annotated. **D)** Principal components (PCs) explaining more than 1% of the variation in NMR metabolite levels between samples (**Methods**). **E)** Spearman correlation coefficients between the PCs and a selection of biological and environmental covariates. Heatmap cells are white and correlation coefficients are dulled where absolute value < 0.1. **F)** Separation of males and females by PC1 and PC3, the two PCs most strongly correlated with sex. The first plot shows hexagonal bins of samples, coloured by sex, with the two plots below showing density plots for PC1 and PC3 stratified by sex.

#### Supplementary Table Legends

**Table S1: Biomarker details.**
Biomarker: variable name used by Nightingale and column name in raw data. Description: full name or description of the biomarker. Units: units of absolute concentration. Group: biomarker group as listed by Nightingale. Sub-group: biomarker sub-group as listed by Nightingale. Type: indicates whether the biomarker is one of the 107 non-derived biomarkers, or can be derived from these 107 non-derived biomarkers. Formulae for biomarker derivation are provided in **Table S2** and code for biomarker derivation is available in the ukbnmr R package (**Code Availability**). UKB Field ID: field ID for the biomarker in UK Biobank. QC Flag Field ID: field ID for the corresponding biomarker measurement QC flags in UK Biobank. Biomarkers without UKB Field IDs are the 76 additional derived biomarkers that are not part of the Nightingale platform and not available to download from UK Biobank.

**Table S2: Derivation formulae for derived biomarkers.**
Formula for deriving each of the 142 composite biomarkers, ratios, and percentages on the Nightingale platform as well as for the 76 additional derived biomarkers. For each biomarker two formulae are given: a formula that uses only the 107 non-derived biomarkers, and a simplified formula which expresses each biomarker in terms of its most relevant parts, in many cases using other derived biomarkers. Code for biomarker derivation is available in the ukbnmr R package (**Code Availability**).

**Table S3: Offsets for log transformation of biomarkers with concentrations of 0.**
Log transformation cannot be applied to values of 0. For these biomarkers, a small offset was added to all concentrations so that all values were greater than 0. The offset used for each biomarker was half of the lowest non-zero concentration. When returning technical covariate adjusted residuals to absolute concentrations (**Figure S8**), after inversing the log transformation and removing the log offset, a small offset was required to prevent negative values. Note that these are one or more orders of magnitude smaller than the minimum non-zero value.

**Table S4: Annotations of GWAS peaks for alanine and histidine.**
Lead SNPs and gene annotations for each independent LD block (**Methods**) with $P < 5\times10^{-8}$ for any variant in GWAS of post-QC alanine and histidine concentrations. Genomic locations are on genome build GRCh37. Beta indicates standard deviation change in quantile-normalized log biomarker concentrations per copy of the effect allele, and SE the standard error. The frequency corresponds to the effect allele frequency in the UK Biobank participants analysed (N=111,743 for alanine, N=111,575 for histidine, **Methods**). The closest protein coding gene for the lead SNP (**Methods**). Where the lead SNP was located in the gene, the most severe consequence and variant impact from the Ensembl variant effect predictor (VEP) are reported, along with variant effect predictions from PolyPhen-2 and SIFT (**Methods**).

**Table S5: Associations between biomarkers and incident coronary artery disease and stroke**
Hazard ratios (HR), 95% confidence intervals (L95, U95), and P-values from Cox proportional hazards models fit on raw and post-QC biomarker concentrations for incident coronary artery disease and incident stroke over 12.8 years of follow-up (**Methods**). Models were fit adjusting for age and sex. Participants with prevalent events or taking lipid lowering medication were excluded. Biomarker concentrations were log transformed (logit for percentages) and standardised for model fitting: hazard ratios are per standard deviation increase in log biomarker concentrations / logit biomarker percentages. Biomarkers are sorted by P-value for association between post-QC concentrations and coronary artery disease.

### Supplementary Notes

***Unsuccessful approaches to removing technical variation***

Several additional approaches to identifying and removing technical variation were explored without success. In particular, we attempted to utilise both the internal control samples and blind duplicate participants to estimate and remove inter-plate variation.

On each plate, internal controls were measured on opposite corners in wells A01 and H12 (**Methods**). These internal control samples were measured in pairs, with a "high" control sample in well A01 and a "low" control sample in well H12 (**Supplementary Methods**). In total, four pairs of internal controls were measured across all plates (**Supplementary Methods**). Theoretically, concentrations for each biomarker should be identical across plates for each control sample. Based on this, we attempted to use Removal of Unwanted Variation (RUV) K-means (De Livera et al., 2015) to learn and remove the inter-plate variation based on the differences between concentrations across plates within each set of control sample pairs. However, inspection of post-QC plots (similar to those shown in **Figure 3G-I** and **Figure S3G**) showed no reduction in inter-plate variation while also showing large structural changes in concentrations when comparing raw to post-QC values (data not shown). We also attempted a simpler approach of estimating inter-plate variation from the internal control samples as their difference from the median across all plates they were measured on, with similar results (data not shown). Both approaches were also explored for identifying outlier plates in the post-QC data, with similar unsuccessful results.

We also explored the possibility of using the blind duplicate samples (**Methods**) to estimate inter-plate variation. However, there were several complications that led us to abandoning such an approach. First, there were 5 plates containing no blind duplicate samples. Second, it was unclear how to utilize the blind duplicates as each were only measured twice (or in a few cases three times), leading to each plate having a mixture of different blind duplicates. Most plates (N=1,282) had four samples belonging to blind duplicates, and a small number (N=18) comprised almost entirely of blind duplicate samples. Additionally there were several participants (N=6) with blind duplicate samples where the multiple measurements were taken on the same plate.

Finally, we also explored adjusting biomarker concentrations for the various sample and biomarker quality control tags provided by Nightingale, for example tags indicating sample dilution. These also had the effect of introducing large structural changes when comparing raw to post-QC values (e.g. shifting all Albumin levels below 30 g/L up by 10 g/L; data not shown).

#### Supplementary Methods

##### *Sample quality control of pre-release data*

Pre-release raw data was provided to early access analysts in a single flat file of wide format with 126,846 rows: with each row corresponding to a single well on a 96 well-plate and columns corresponding to the 249 quantified biomarkers along with sample information.

First, on the advice of Nightingale Health Plc., we removed from the raw data 40 rows corresponding to samples erroneously measured despite having insufficient sample material. These included 37 samples present in the raw data available to download from UK Biobank (**Methods**). Next, 441 rows with missing values for all 249 biomarkers were removed.

Next we resolved duplicate samples. In total, there were four samples measured twice (rows with the same Nightingale sample identifier and UK Biobank visit annotation, measured on different plates and wells): samples with Nightingale identifiers 1219449, 4478891, 2553581 and 1556057, each corresponding to an anonymised UK Biobank participant at baseline assessment. From each pair of duplicates one sample was kept. For samples with Nightingale identifiers 1219449 and 4478891, the rows corresponding to measurements on plate 490000006107 were removed as the sample annotation information indicated these were measured on spectrometer 10176949, whereas the other 92 samples on that plate were measured on spectrometer 10278626. For samples with Nightingale identifiers 2553581 and 1556057, the rows corresponding to measurements on plate 490000006069 were removed as they were reportedly measured 48 hours prior to the rest of the samples on plate 490000006069, whereas their measurements on plate 490000006068 occurred day as all other samples on that plate.

Finally, samples and biomarker measurements with quality control tag “technical error” were resolved. Nightingale Health Plc. advised that all samples and biomarker measurements with this tag should be set to missing. Among the 189 samples with this tag, one sample had non-missing data and was removed (the other 188 were removed above as they already had all missing data). Among the 2,392 biomarker measurements with this tag (across 292 samples), 69 measurements (in 3 samples) were not missing in the raw data and subsequently set to missing here. All samples with this quality control tag have data set to missing in the raw data available to download from UK Biobank.

In total 126,360 samples passed quality control. These included 6,359 blind duplicate samples: samples from participants sent by UK Biobank multiple times with differing sample identifiers for UK Biobank to assess the internal consistency of Nightingale Health’s NMR metabolite biomarker quantification pipeline. In total, 121,758 participants passed quality control: 118,047 with measurements at baseline assessment and 5,139 with measurements at first repeat assessment, including 1,428 participants with measurements at both timepoints.

##### *Quality control of internal control samples*

Internal control samples were placed on opposite corners of each of the 1,352 plates (wells A01 and H12) by Nightingale Health to assess internal consistency of their NMR metabolite biomarker quantification pipeline. Data from these internal control samples was provided to early access analysts in a single flat file of wide format with 2,713 rows: with each row corresponding to a single well on a 96 well-plate and columns corresponding to the 249 quantified biomarkers along with sample information. In total, 2,698 control samples passed quality control.

First, duplicate internal control samples resolved. For plate 490000006069, there were two rows corresponding to wells A01 and H12, each pair of duplicates with internal control identifiers 180827 and 180829 respectively. From each pair, we kept the measurement taken closest in time to the rest of the plate. For plate 490000006107, there were two rows corresponding to wells A01 and H12, each pair of duplicates with internal control identifiers 180829 and 180827 respectively. From each pair, we kept the measurement taken on the same spectrometer (10278626) as the rest of the plate.

Next, several other problematic internal control samples were also identified and removed. Control samples measured in well H12 on plates 490000007201 and 490000006663 were discarded, as a “high” control sample was used instead of a “low” control sample: control sample 190404 was measured in well H12 on these two plates but in well A01 on 213 other plates. Internal control samples 180817, 190125, and 191127 were also removed as they were measured on only 1-2 plates each in well A01, whereas the other 8 internal control samples were measured in pairs on 213–444 plates each.

Finally, four failed internal control samples with extreme biomarker concentrations were removed on the advice of Nightingale Health Plc: control sample 180827 on plate 490000006201 in well H12, control sample 180829 on plate 490000005965 in well A01, control sample 190328 on plate 490000006714 in well H12, and control sample 190328 on plate 490000006802 in well H12. None of these four plates were flagged as outliers for any biomarker when identifying outlier plates driven by unexplained technical variation (**Methods**).
